# Routine respiratory metagenomics service for intensive care unit patients

**DOI:** 10.1101/2023.05.15.23289731

**Authors:** Themoula Charalampous, Adela Alcolea-Medina, Luke B. Snell, Christopher Alder, Mark Tan, Tom G.S. Williams, Noor Al-Yaakoubi, Gul Humayun, Christopher I.S. Meadows, Duncan L.A. Wyncoll, Paul Richard, Carolyn J. Hemsley, Dakshika Jeyaratnam, William Newsholme, Simon Goldenberg, Amita Patel, Fearghal Tucker, Gaia Nebbia, Mark Wilks, Meera Chand, Penelope R. Cliff, Rahul Batra, Justin O’Grady, Nicholas A. Barrett, Jonathan D. Edgeworth

**Author notes:** These authors contributed equally (Themoula Charalampous, Adela Alcolea-Medina, Luke B. Snell). Senior author (Jonathan D. Edgeworth). Corresponding author for respiratory metagenomics and for clinical.

## Abstract

**Background:** Respiratory metagenomics (RMg) needs evaluation in a pilot service setting to determine utility and inform implementation into clinical practice.

**Methods:** Feasibility, performance and clinical impacts on antimicrobial prescribing and infection control were recorded during a pilot RMg service for patients with suspected lower respiratory tract infection (LRTI) on two general and one specialist respiratory intensive care units (ICU) at Guy’s & St Thomas NHS foundation Trust, London.

**Results:** RMg was performed on 128 samples from 87 patients during the first 15-weeks providing same-day results for 110 samples (86%) with median turnaround time of 6.7hrs (IQR 6.1-7.5 hrs). RMg was 92% sensitive and 82% specific for clinically-relevant pathogens compared with routine testing. 48% of RMg results informed antimicrobial prescribing changes (22% escalation; 26% de-escalation) with escalation based on speciation in 20/24 cases and detection of acquired-resistance genes in 4/24 cases. Fastidious or unexpected organisms were reported in 21 samples including anaerobes (n=12), *Mycobacterium tuberculosis, Tropheryma whipplei*, cytomegalovirus and *Legionella pneumophila* ST1326, which was subsequently isolated from the bed-side water outlet. Application to consecutive severe community-acquired LRTI cases identified *Staphylococcus aureus* (two with *SCCmec* and three with *luk* F/S virulence determinants), *Streptococcus pyogenes* (*emm1-*M1uk clone), *S. dysgalactiae* subspecies equisimilis (STG62647A) and *Aspergillus fumigatus* with multiple treatments and public-health impacts.

**Conclusions:** RMg provides frequent diverse benefits for treatment, infection control and public health. The combination of rapid comprehensive results, alongside revealing and characterising a hidden burden of infections makes the case for expediting routine service implementation.

## Introduction

Community and hospital acquired lower-respiratory tract infections (LRTI) are caused by an expanding range of mono-microbial and polymicrobial infections. This presents significant challenges identifying or excluding microbial cause(s). Typically, samples are tested by different methodologies with results returned at different times over subsequent days (1). Culture remains the gold-standard for bacterial identification, despite being slow, having suboptimal sensitivity particularly after antibiotic treatment and inability to detect many fastidious organisms. Multiplex PCR-testing detects many pathogens and can add value to early decision-making, but they target a restricted repertoire, do not provide genomic detail and cannot exclude infection.

RMg has potential to become a first-line test for severe pneumonia, given its ability to identify essentially any microbe in a clinical sample along with antimicrobial resistance and virulence determinants (2). Retrospective and prospective proof-of-concept studies and case series have been published (3-8) but none have demonstrated feasibility and the breadth of impact from a single test as a routine daily service. We developed a 6-hour nanopore sequencing workflow (6) and evaluated its potential during the COVID-19 pandemic (2). Here we present results implementing RMg into pilot service for ventilated ICU patients.

## Methods

### Setting and sample collection

RMg testing was offered to a 41-bed medical, surgical and specialist respiratory ICU that included an Extracorporeal Membrane Oxygenation (ECMO) service, Monday to Friday between November 22^nd^-December 15^th^ 2021 and January 4^th^-March 25^th^ 2022. Pilot service provision was agreed by the Critical Care Governance & Audit Committee under the NHS Quality Improvement and Patient Safety (QIPS) governance process as previously described (9) (QIPS reference 2021:13023). Duty intensivists selected mechanically ventilated (MV) patients to have additional RMg testing alongside culture and 16S rRNA sequencing performed by the clinical laboratory. Decision criteria were based on the potential for a rapid result to assist with diagnosis or exclusion of LRTI and antibiotic prescribing decisions. Respiratory samples were retrieved prospectively from ICU at 8.30am with results aimed before 5pm. Representative severe CA-LRTI cases from the first 8 weeks of the following influenza season (winter 2022) are also presented, which were absent from the pilot conducted during the COVID-19 pandemic.

### Respiratory metagenomic sequencing workflow

In total 128 samples were processed for RMg sequencing which included 111 bronchoalveolar lavages (BAL), 3 tracheal aspirates, 8 non-direct bronchoalveolar lavages (NDL) and 6 pleural fluids. RMg testing involved saponin-based host depletion, microbial extraction, library preparation and nanopore sequencing as previously described (2, 6, 10) (Figure 1). Every RMg run also included quality controls (no template control (NTC), positive control (PC) and a competitive spiked-in internal control (IC) to identify run or single-sample failures and contamination. Method detailed in supplementary material.

**Figure 1.**
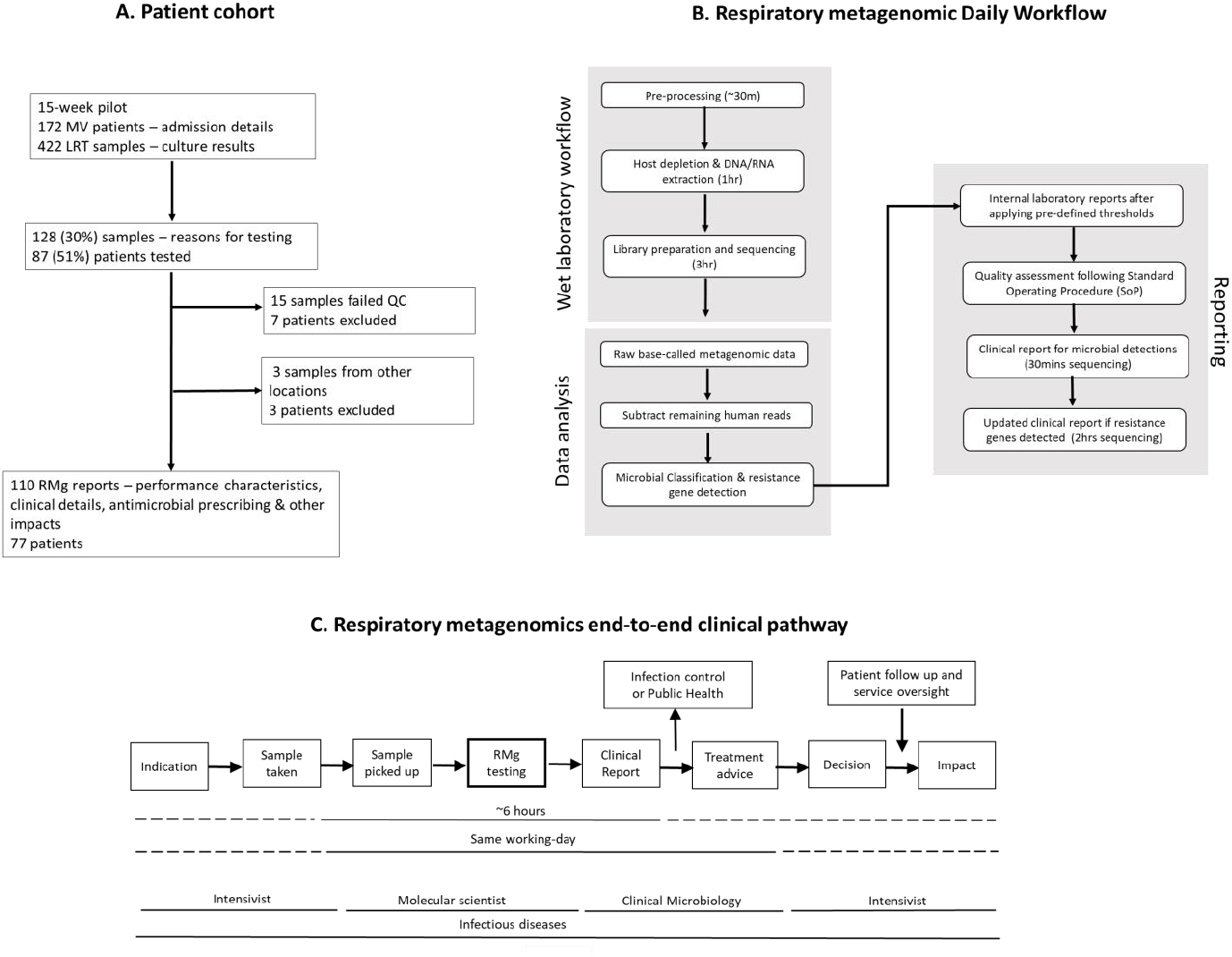
Schematic overview of the study. Overview of the patient cohort and sample set included in the RMg pilot service (A). The metagenomics regime followed on a daily basis when samples were requested for RMg service. Steps outlined include sample collection until reporting results to ICU physicians (B). Respiratory metagenomic end-to-end clinical pathway (C). MV= Mechanically-ventilated; LRT= lower respiratory tract; RMg= Respiratory metagenomics; QC= quality control.

Time-point data analysis was performed using an in-house pipeline (https://github.com/GSTT-CIDR/RespiratoryCmg) incorporating taxonomic classification, antimicrobial resistance (AMR) gene detection and sequence-based typing (SBT). The k-mer based classifier Centrifuge (11), was used for microbial identification using the FDA-ARGOS database (12), curated with only respiratory organisms containing 673 microbial sequences.

#### Reporting of RMg results

Sequencing reports generated at 30 minutes and 2 hours were interpreted following the Standard Operating Procedure (SOP). Bacteria were reported using 30 minutes sequence-data when representing ≥1% total microbial reads and fungi if ≥5 reads were detected, and with Centrifuge score ≥8000. Microorganisms reported from the RMg workflow in this study were referred to as ‘respiratory pathogens’ or ‘pathogens’ and they were defined as microorganisms causing respiratory infection in ICU patients. A list of reportable organisms was compiled and followed for reporting. The list was based on previous LRTI studies (1, 2, 12-14) and previous findings from the archives of microbiological culture in the last 5 years collected from the clinical laboratory (Table S10). Organism-specific reporting criteria were also set for certain organisms (such as *E. faecium* and anaerobes) - reporting is described in detail in supplementary material.

Acquired resistance genes reporting restricted to extended-spectrum beta lactamases in Enterobacterales, SCCmec in Staphylococcus aureus and Vancomycin-resistance gene clusters in *Enterococcus faecium* were included in 2 hour sequence reports (2).

Clinical sequence reports were uploaded as a pdf-file to the ICU electronic health record after scientific and medical review. Results were also communicated by email or verbally to the duty intensivist and infectious diseases doctor (Figure 1C). RMg performance was compared with routinely requested tests. Discrepant results were investigated using routine 16S rRNA sequencing testing or an in-house pathogen-specific targeted qPCR.

### Data Availability

Sequencing data presented in this study are available to the European Nucleotide Archive (ENA) under project number PRJEB59568.

### Funding

This work was supported by the National Institute for Health Research (NIHR) Biomedical Research Centre programme of Infection and Immunity (RJ112/N027), Guy’s & St. Thomas’ Charity (https://www.gsttcharity.org.uk/; TR130505) and the Medical Research Council (MR/W025140/1; MR/T005416/1 for LBS and GN) and (MC_PC_19041).

### Patient management and impacts of RMg

Clinical, microbiology and antimicrobial prescribing data were collected prospectively from all MV-ICU patients having at least one LRT sample collected during the pilot, alongside RMg. Samples from patients in non-pilot critical care areas were excluded for downstream analysis and data colelction. RMg-based antimicrobial treatment changes and findings of infection control or public health importance communicated by email or phone calls the same day. RMg results and impacts were reviewed biweekly by multidisciplinary team of three intensivists, an infectious disease doctor, two microbiologists and a pharmacist. Clinical implications of unexpected or discrepant results or any adverse impacts of interventions in response to RMg results were reviewed.

## Results

### Clinical and microbiological characteristics of ventilated ICU patients

172 ventilated patients admitted to ICU during the 15-week period had 422 LRT samples cultured. In total 128/422 (30%) LRT samples from 87/172 (51%) patients had additional RMg testing (Figure 1A). Clinical characteristics of RMg tested patients was similar to non-tested patients apart from more COVID-19 infections (33% vs 21%) and ECMO therapy (38% vs 5%). (Table 1). 9/172 (5%) patients had any LRT sample growing Gram-negative bacteria (GNB) phenotypically-resistant to first-line empiric treatment of hospital-acquired LRTI (piperacillin-tazobactam) and 3 patients had vancomycin-resistant *E. faecium* (VRE) or carbapenem resistant *P. aeruginosa*. No cases of MRSA or carbapenem-resistant Enterobacterales were reported (Table S1).

**Table 1.**
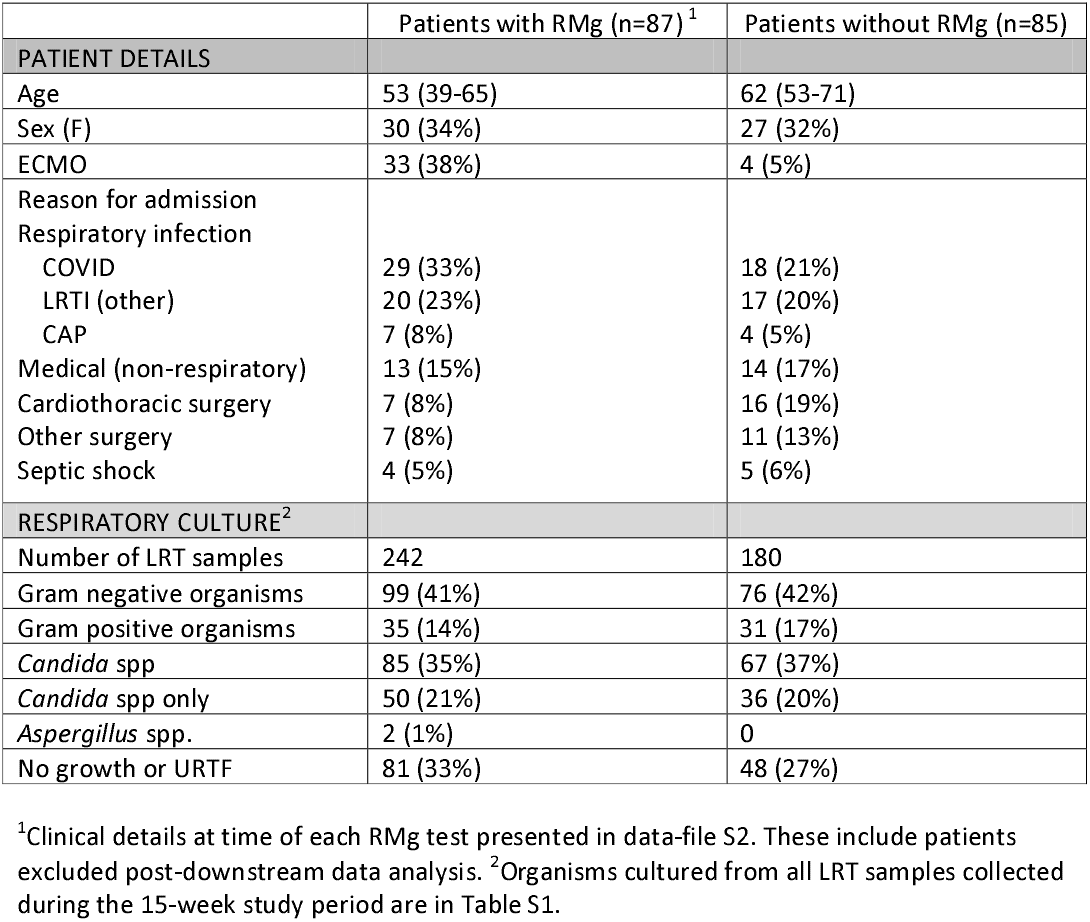
Clinical characteristics and routine microbiological testing of the patient cohort during 2021-2022 winter seasons.

### RMg performance against routine testing

RMg was performed on samples sent for new suspected community-acquired (CA)-LRTI (23%), at the start (29%) or during (34%) an episode of suspected hospital-acquired (HA)-LRTI or for other reasons (14%) (Table 2). These included 15/128 samples failed QC ((n=7 patients (Table S2)) and 3/128 samples were also excluded due to patients (n=3) admitted to different acute wards. These samples were excluded from further analysis and performance of RMg was calculated using the remaining samples (n=110). The median TAT from sample receipt to RMg reporting was 6.7hrs (interquartile range 6.1-7.5 hrs: maximum 30.5 hrs), with 90% having same-day final reports. This compared with verbal communication of interim culture results when available the following afternoon (median 29hrs) and final reports generated at median of 40hrs.

**Table 2.** Antimicrobial treatment changes in response to RMg results.

| Indication for RMg testing | N=128 |
| --- | --- |
| CA-LRTI | 30 (23%) |
| HA- LRTI | 80 (63%) |
| - Start of episode | 37 (29%) |
| - During episode | 43 (34%) |
| - Other<sup>1</sup> | 18 (14%) |
| Antibiotic prescribing<sup>2</sup> | N=110<sup>3</sup> |
| Receiving antibiotics start of RMg test day | 89 (81%) |
| Receiving antibiotics end of RMg test day | 98 (89%) |
| De-escalation: antibiotics stopped with RMg<sup>4</sup> | 29 |
| - Same day | 7 (24%) |
| - Next day<sup>5</sup> | 22 (76%) |
| Meropenem | 15 |
| Piperacillin-Tazobactam | 6 |
| Linezolid | 5 |
| Other | 3 |
| Escalation: antibiotics started with RMg<sup>6</sup> | 24 |
| - Same day | 21 (87%) |
| - Next day<sup>7</sup> | 3 (13%) |
| Meropenem | 10 |
| Linezolid | 4 |
| Other | 10 |

<sup>1</sup> Includes where the focus of infection was uncertain at time of testing, to exclude respiratory infection pre-immunomodulation or after completing therapy for LRTI. <sup>2</sup>15 patients had antibiotics started or stopped for reasons not linked with the RMg result.
<sup>3</sup>Excludes samples failed at QC (n=15) or sent from non-pilot critical care areas (n=3).
<sup>4</sup>Antifungals were stopped in 3 patients in response to RMg results <sup>5</sup>2 patients had antibiotics stopped on second day after testing. <sup>6</sup>Antifungals were started for 5 patients in response to RMg results <sup>7</sup>3 patients had antibiotics started in response to RMg results two days after receipt of the result

In 44/48 culture-positive samples RMg was in agreement with culture findings. RMg missed culture-reported organisms in 4/48 samples. All missed organisms were reported as scanty growth by culture (*S. aureus* [P2 and P77] and *K. pneumoniae* [P7 and P9] (Figure S1). RMg did not detect clinically-relevant organisms in 51/62 samples reported as ‘negative for pathogens’ by culture. These included 25/51 samples reported as ‘negative’ or ‘no growth’ by culture and 26/51 samples reported positive for commensal or non-clinically-relevant organisms (such as *Candida* spp.) (Figure S1). Clinically-relevant organisms were detected by RMg in 11/62 samples – these included *S. aureus* (n=3), *E. faecium* (n=2), anaerobic bacteria (n=4), *<u>C. striatum</u>* (n=1) and *C. koseri* (n=1).

Based on these findings RMg was 92% (95% CI, 80-98%) sensitive and 82% (95% CI, 71%-91%) specific on a per-sample basis compared with culture. Only 3/11 findings were not confirmed via confirmatory testing (Table S3) which increased specificity of RMg to 94% (95% CI, 85-99%). The increase in specificity after including confirmatory testing was mostly due to RMg identifying bacteria at low levels and organisms subsequently identified in other samples types in the same patients (Table S3). Overall RMg reported organisms not reported by routine testing in 26/110 (17%) samples of which 15/26 were culture-positive samples. These included DNA viruses (HSV-1 (n=5) and CMV (n=1)) and bacteria ((n=9) (data file S1)). Additional RMg findings in culture-positive samples were not considered false-positive findings.

RMg findings were also compared against findings from routine 16S rRNA sequencing. In total 85/110 RMg samples were also send for additional 16S rRNA sequencing by the clinical laboratory from which 50/85 (59%) samples were concordant with RMg findings. From the remaining samples 25/85 (29%) were discordant and 10/85 (12%) samples both test were in agreement for ≥1 microbial detection.

Acquired resistance genes were also reported in 5 samples, vanA [P88], blaSHV [P96, P115 and P122] and bla*CXT-M* [P117]. RMg missed *K. pneumoniae* bla*CXT-M* in one patient [Figure 2 patient D and sample P110) but it was identified by repeat sequencing using MinION flow-cell that has higher sequencing yields (13)(data-file S1).

**Figure 2.**
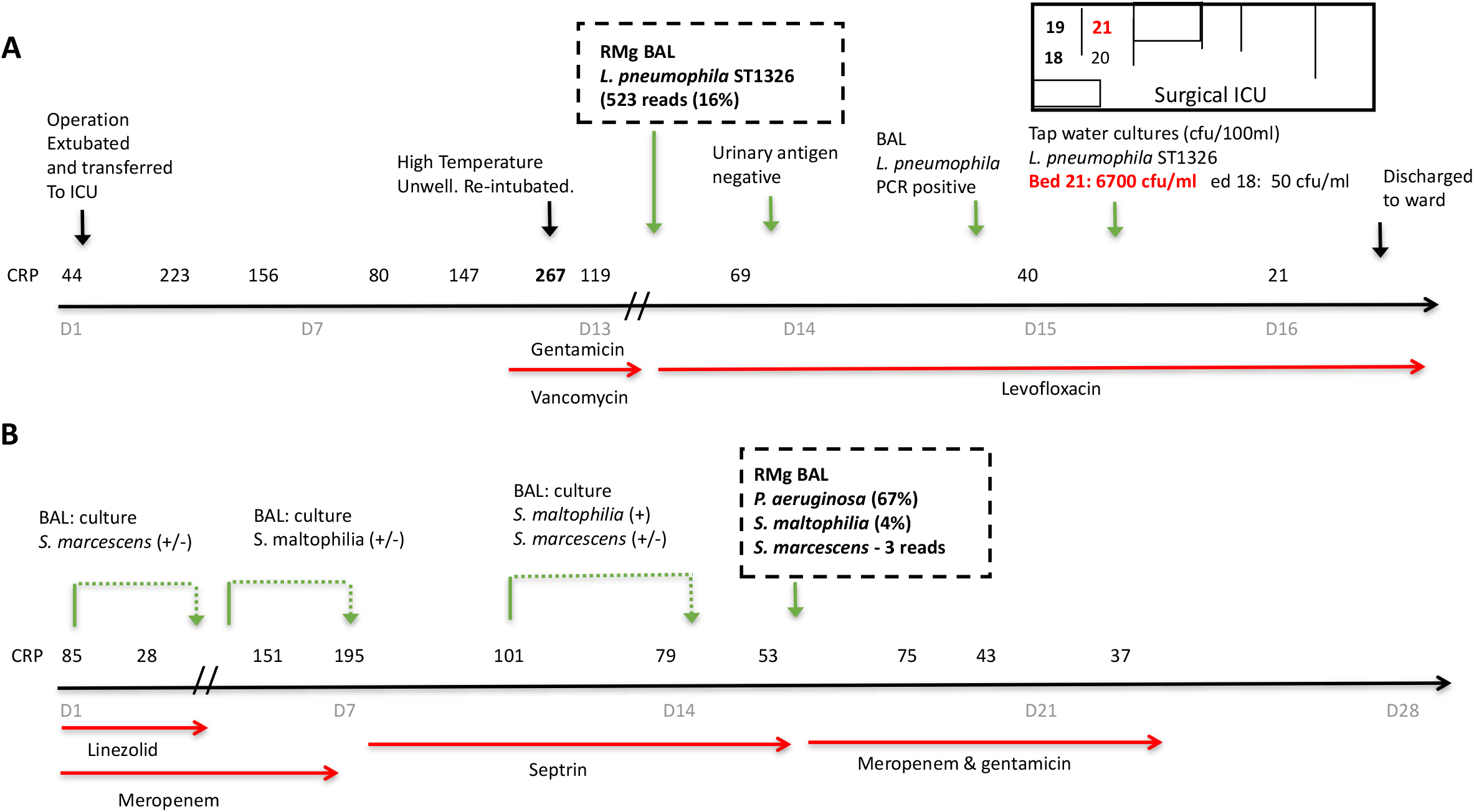
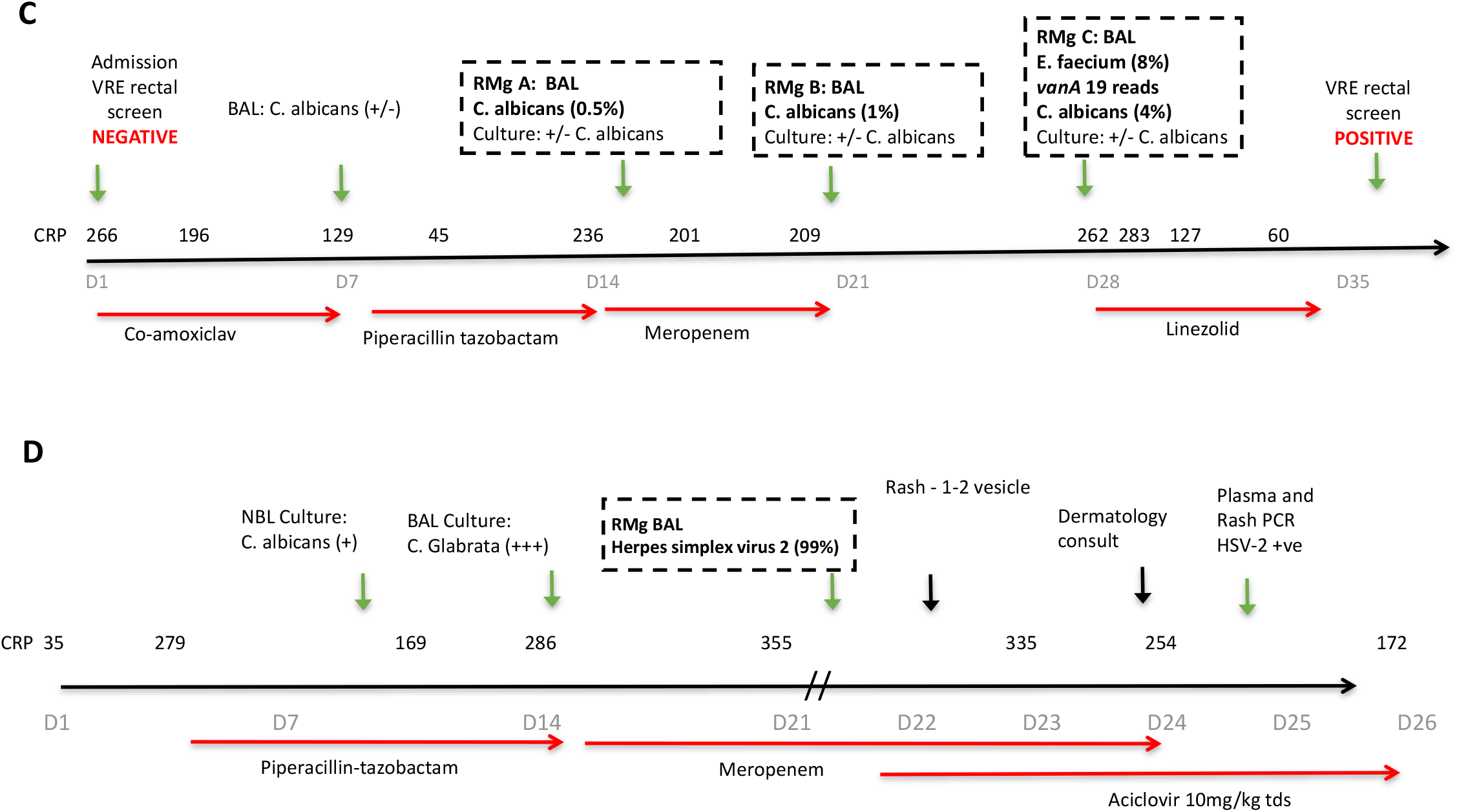
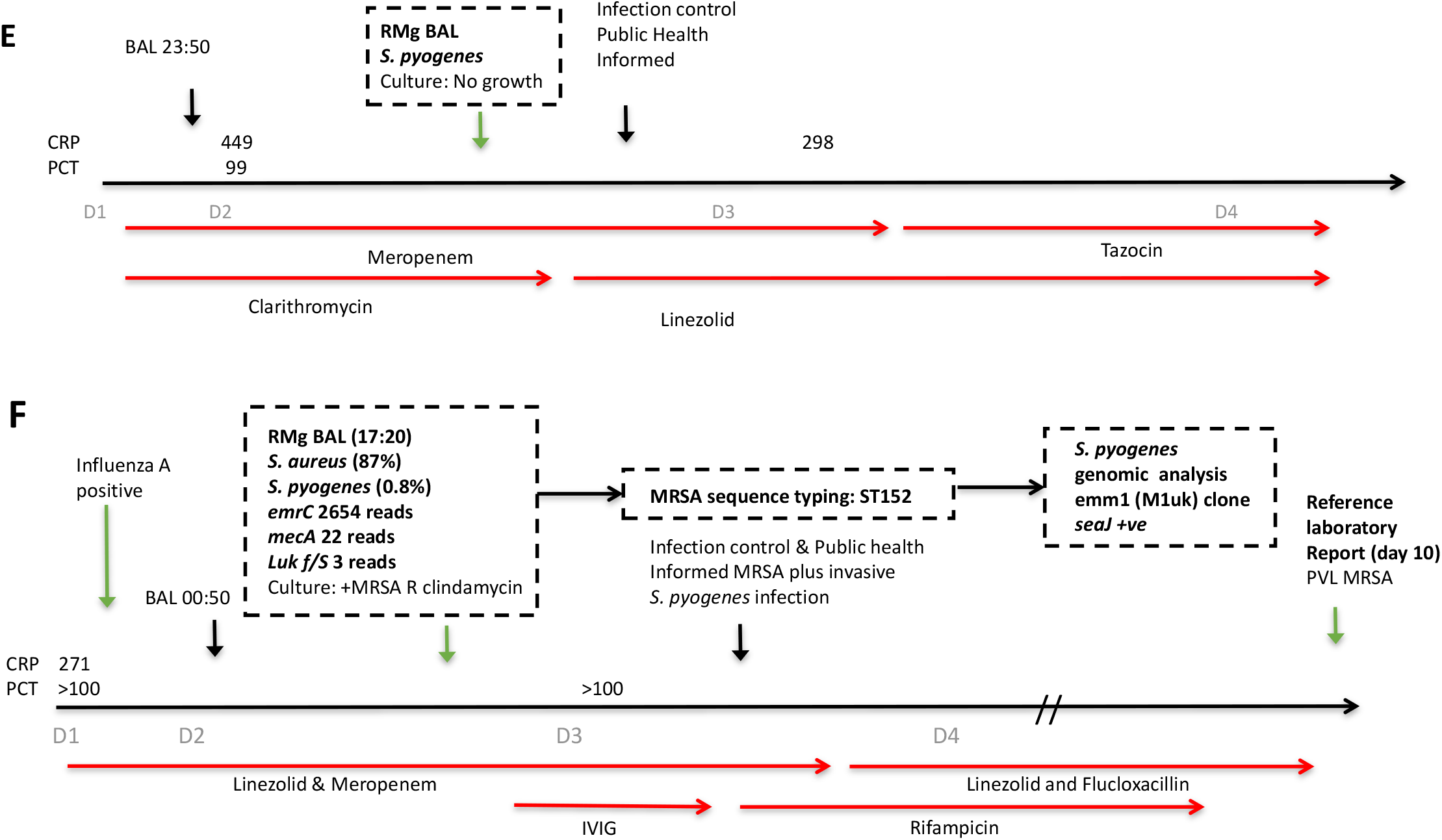
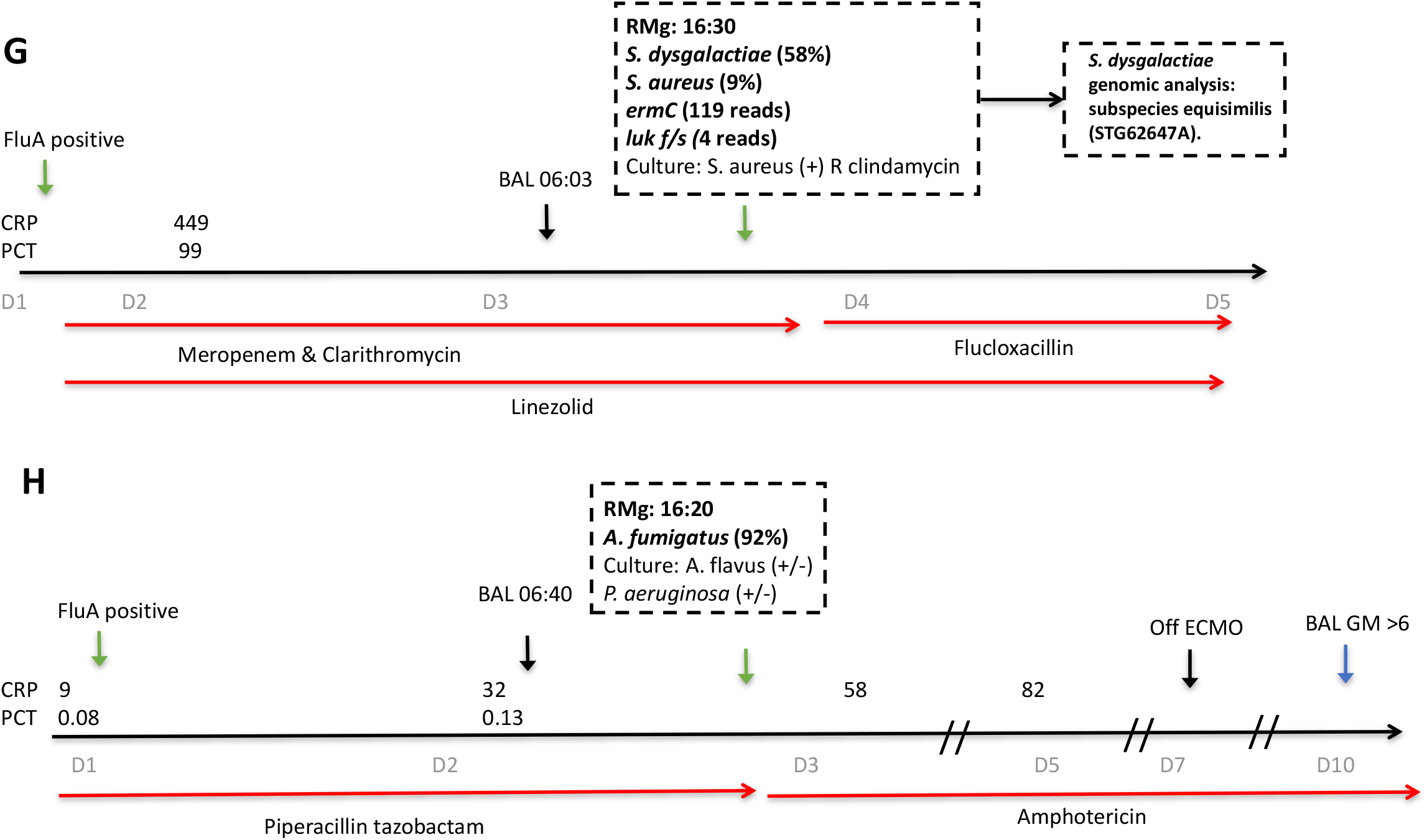
Patient ICU-timelines illustrating integration of RMg results into antimicrobial treatment and infection control decisions. Hospital-acquired LRTI A) ICU acquired *L. pneumophila* ST1326 pneumonia. Unexpected bacteria prompting antibiotic escalation, infection control and public health interventions. B) *<u>P. aeruginosa</u>* VAP-LRTI. New bacterial pathogen in patient with severe COVID-19 pneumonitis prompting antibiotic escalation. C) ICU-acquired vancomycin resistant *E. faecium*. Unexpected AMR-bacteria with patient & infection control impact. D) Unexpected disseminated reactivation of HSV-2. Community-acquired LRTI E) Influenza with secondary S. pyogenes infection F) Influenza with secondary PVL-MRSA and *S. pyogenes* infection. G) Influenza with secondary PVL-MSSA and *S. dysgalactiae* infection. H) Influenza with secondary invasive aspergillosis prompting urgent treatment. Details or each case are presented in supplementary material.

### Impacts on antibiotic treatment

RMg contributed to treatment decisions in 88/110 (80%) cases (Table 2). In 24 (22%) cases, antibiotics were started (n=10) or escalated (n=14) based on detecting organisms with intrinsic (n=20) or acquired resistance (n=4) to current therapy, the majority (87%) that day (Table 2). In 29 (26%) cases, antibiotics were de-escalated or stopped predominantly the following morning ward round (n=22/29 cases (76%)). De-escalation occurred in 66% (19/29) of cases when RMg detected no clinically-relevant organisms (13 no organisms, 3 upper respiratory tract commensals, 3 *Candida spp*). All de-escalation cases were followed up. One patient had antibiotics re-started to treat *P. aeruginosa* cultured from a respiratory sample taken 3 days after RMg informed stopping of antibiotics [P78]. The patient was otherwise progressing well and left ICU few days later. Clinical details of all cases are in data-file S2, with representative timelines for 3 HA-LRTI cases in Figure 2 cases A-C.

For 35 (32%) patients, antimicrobials were not changed but contributions to prescribing decisions were recorded, mostly by reassuring clinicians of no unexpected pathogens. Thereby preventing escalation, particularly in heavily immunosuppressed patients and with persistent inflammation on antimicrobial treatment (n=11). RMg results also prompted early immunomodulation for suspected inflammatory lung conditions (n=7) after excluding pathogens (Table 3).

**Table 3.** Categorising impact of respiratory metagenomic results on antibiotic prescribing.

| Treatment category | Organisms identified by RMg<sup>1</sup> | Antibiotic changes |
| --- | --- | --- |
| START OR ESCALATE TO ACTIVE ANTIMICROBIALS N=24 [22%] | AmpC Enterobacterales (9); Anaerobes (3); *Candida* spp (2); *P. aeruginosa* (3); VRE (1); ESBL *K. pneumoniae* (1); *H. influenza* and *K. variicola* (1)<sup>2</sup>; *H. influenza* and *M. morganii* (1); *E. faecium* (2); *C. striatum* (1) | STARTED: Meropenem (10); Linezolid (3); Linezolid & Anidulafungin (1); Ciprofloxacin (1); Piperacillin-Tazobactam (2); Ciprofloxacin (1); Other (6) |
| DE-ESCALATE OR STOP N=29 [26%] | No significant organisms (29) (comprising no organisms (13), *Candida* spp (3), URTF (3) Anaerobes (3); MSSA (2); *K. variicola* & *H. influenzae* (3); *E. faecalis* (1); *E. coli* (1) | STOPPED: Meropenem (6); Piperacillin-Tazobactam (4); Piperacillin-Tazobactam & Gentamicin (2); Linezolid (1); Meropenem & Linezolid (4); Meropenem & Other (5); Co-amoxiclav (1); Ciprofloxacin (1); Levofloxacin (2); Other (3) |
| PREVENT ESCALATION, REASSURE OR INFORM OTHER THERAPY N=35 [32%] | No significant organisms: No organisms (10), *Candida* spp only (3), URTF (2); Anaerobes (6); *K. pneumoniae* (3); Other Enterobacterales (4); MSSA (4); *Candida* spp. [with MSSA] (1), *C. striatum* (1), *E. faecium* (1) | Reassure correct antimicrobial chosen (17); Exclude organisms to bring forward immunomodulation (7). Exclude untreated organisms in complex or immunosuppressed patients (8) Persistent inflammation on current antibiotics (3) |
| NO TREATMENT IMPACT N=22 [20%] | No significant organisms: No organisms (11), *C. albicans* (2), *K. pneumoniae* (3), other Enterobacterales (3) and 1 each of *P. aeruginosa*, *E. faecalis*, MSSA (2) | Clinical concern prevented de-escalation in response to RMg result (6); Potential or proven infection at other site (8); Delay returning result (4); Missed information [resistance] (2); quantity of organism (1) organism missed (1) |

Impact categories were: I) earlier appropriate antimicrobials where the result prompted changes to existing therapy to target the identified organism(s), II) de-escalating antimicrobials where the result contributed to stopping or narrowing antimicrobial spectrum III) prevent antimicrobial escalation, reassuring that current therapy was appropriate or informing non-antimicrobial treatment therapy and IV) no identified benefit for range of reasons.
<sup>1</sup>Organisms identified by RMg in monomicrobial and polymicrobial LRT samples. <sup>2</sup>*K. variicola* and *H. influenza* grown but susceptibilities not available. *H. influenza* resistant to coamoxiclav but patient improving and extubated so patient completed 5 day course. Coamoxiclav started 2 days later. ESBL= Extended spectrum b-lactamases, MSSA=methicillin-sensitive *S. aureus*, URTF= Upper respiratory tract flora. VRE= Vancomycin-resistant *E. faecium*

Anaerobes detected in 12 samples (10 BALs and 2 PFs) were deemed clinically-relevant based on clinical findings and absence of alternative plausible pathogens. Patients with CA-LRTI (n=8) had history of aspiration and patients with HA-LRTI (n=4) had received antimicrobials for between 5-12 days, some of which lacked anaerobic activity. Antibiotics were started (n=4), de-escalated (n=3) or continued (n=5). Conversely their exclusion from a clinically-suspected lung abscess and empyema prompted a diagnosis of lung infarction with hydro-pneumothorax and shortening of planned antibiotic course from 6 weeks to 5 days (Table S4).

22/110 (20%) RMg findings had no recordable impact either because infection was diagnosed at another site (n=8), results were not acted upon (n=6) or decisions were made before results were returned (n=4) (Table 3).

### Information for infection control

Same-day communication of one VRE and two ESBL cases prompted early institution of barrier precautions. The VRE (P88 and Figure 2C) was only cultured from a rectal swab requested during follow up, in response to RMg results. Additionally, *K. variicola* (3 patients) transmission based on overlapping ward-stays was investigated using RMg data. *K. variicola* from 2/3 patients had 99.9999% genetic similarity implicating patient-to-patient transmission (genetic similarity also confirmed by whole genome sequencing of the isolate).

### Unexpected organisms reported by RMg

RMg reported organisms not generally detectable by tests requested during initial patient investigation in 9 samples that were classified as unexpected. *L. pneumophila* ST1326 (serogroup 10) was an ICU-acquired infection 13 days post-cardiothoracic surgery [P123], confirmed by PCR but not urinary antigen testing. The same sequence type was isolated from the adjacent hand-basin tap water (Figure 2 case A), so new water filters were fitted to prevent further cases. *M. tuberculosis* was detected in a patient admitted with haemoptysis few months after starting anti-tuberculous therapy that was interpret as dead organisms so no action was taken. That sample and 4 further samples were auramine and culture negative. *Tropheryma whipplei* [P103] was detected in a patient with HA-LRTI post thymoma resection prompting ceftriaxone treatment and follow up by infectious diseases but the significance remained uncertain. CMV detection in a patient with Jo-1 anti-synthetase deficiency prompted plasma viral load testing which was positive (log 3.5-3.6). It was considered clinically-relevant and ganciclovir was commenced [P35]. HSV-1 was detected in 5 samples, but all were considered non-pathogenic reactivation.

### Representative cases from 2021/2022 winter season

8 severe CA-LRTI cases were admitted over an 8 week period of which 6 were co-infections with influenza (Table S5A). RMg identified Panton Valentine Leukocin (PVL)-*S. aureus* (3), *S. pyogenes* (2), *S. pneumoniae* (2), *S. dysgalactiae* (1), *L. pneumophila* (1) and *A. fumigatus* (1). Only one streptococcus (*<u>S. pneumoniae</u>*) was cultured. Treatment was escalated in 3 cases that day (addition of linezolid, intravenous immunoglobulin or ambisome (Figure 2E-2H). PVL-*S. aureus* and *S. pyogenes* cases were reported to public health that day. Subsequent analysis of RMg data identified one *S. pyogenes* as *emm1*-M1uk clone and the *S. dysgalactiae* as subspecies *equisimilis* (STG62647A).

16 additional patients had RMg testing of which the most important result was unexpected HSV-2 in a patient with new hepatitis and suspected drug rash post-vascular surgery (Figure 2 case D and table S5). High dose acyclovir was started that day. Subsequent plasma and rash swab samples were HSV-2 positive.

## Discussion

RMg must demonstrate consistent additional clinical benefits beyond current practice to be implemented as a routine first-line test for severe pneumonia on ICU (14, 15). We configured laboratory and clinical pathways into a daily RMg service framework to identify five benefit overlapping categories. Firstly, earlier provision of results than culture usually provides (median 40 hours vs 6.7 hours from RMg) to improve initial antimicrobial treatment. This occurred in almost half of patients and was predominantly due to species identification rather than acquired resistance genes, which were uncommon in this cohort. Secondly, identification of organisms that are hard to identify by culture or are suppressed by prior antibiotics. Anaerobes with or without *S. milleri* were found in 10% of samples, similar to a previous study using the same service framework (9) and have been identified in other RMg studies (7). Anaerobes are a long-recognised (7, 16-18) but overlooked cause of LRTI (19), perhaps due to difficulties with detection. De-escalating antibiotics to target anaerobes could have significant benefit for antimicrobial stewardship given their frequent detection. Thirdly, providing an actionable “negative” result when no (clinically-relevant) organisms are detected, which RMg is uniquely placed to provide. No adverse consequences were identified in cases when de-escalation took place, however safe de-escalation always requires, close monitoring, confidence in sample quality and an understanding of methods limitations and reporting thresholds. This is particularly relevant in acutely unwell patients or when considering immunomodulation for clinically-suspected inflammatory lung conditions. The fourth category was identifying AMR organisms for infection control. One VRE and 2 ESBL cases prompted early institution of barrier precautions. Detection of the VRE was unlikely without RMg testing.

These first four categories represent improvements to the routine culture pathway, but the final category was identifying organisms currently requiring targeted molecular tests that were not requested by intensivists. Some proved clinically-important such as the CMV, HSV-2 and L. pneumophila cases, with the latter prompting interventions to prevent further cases. In contrast, the 5 HSV-1, *M. tuberculosis* and probably the *T. whipplei* cases were not significant; although, all would be in different clinical contexts (20-22). Providing unexpected or un-requested results can prompt unnecessary investigation and treatment; however, identifying benefits in about a third of cases as found here could be considered an acceptable yield.

Utility of this fifth *“molecular”* category is extended when the additional information provided by pathogen sequencing is considered. Targeting the water supply as the source of *L. pneumophila*-ST1326 required sequence-based typing, not just organism identification, as did confirming *K. variicola transmission* (2). RMg applied to severe CA-LRTI cases identified virulence factors (luk f/S) and emerging virulent clones missed by culture (*S. dysgalactiae* STG62647A and *S. pyogenes emm1-*M1uk) (23). The latter is of particular public health significance given its link with severe paediatric infections and deaths, first announced by UKHSA 8 weeks after this case (24, 25). Finally, demonstrating ability to assess genotypic azole resistance in *A. fumigatus* sequence from RMg data would be a significant improvement to current fungal AMR testing.

This study has limitations. Only 3 samples maximum were processed per day due to limited operator availability so the proportionate impact may reduce, when extended to all respiratory samples. Reported sample-failure rate (12%) was mostly due to a defective DNA-extraction batch or operator-introduced contamination which is a recognised current limitation of RMg (26, 27). This can be addressed by reducing hands-on-time via automation. Only phenotypes from acquired-resistance elements were reported, however, expansion to phenotypic prediction caused by other AMR mechanisms (mutational) to increase RMg-usability in settings with higher AMR rates is required. Finally, this workflow is not designed to detect RNA viruses and has not been assessed for other DNA viruses, so workflows that additionally detect these organisms should be evaluated (28).

In conclusion, this pilot service demonstrates the clinical utility of RMg testing in a routine setting reporting organisms usually detected by culture alongside fastidious and/or uncultivable organisms while also providing genomic information for AMR prediction and/or identifying hospital-acquired infections and emerging hyper-virulent community clones aiding infection controls decisions. Realising all these benefits for individual patients and the wider healthcare system will require change to current practice by many professional groups working more closely together in the same acute timeframe (29). Implementation still requires further technology refinement, addressing accreditation and regulatory requirements, along with gathering data from larger multi-centre studies and health-economic analyses (27, 30). Nevertheless, given recognised gaps in preparedness highlighted by the COVID-19 pandemic (31-33) and increasing AMR (34, 35), this study gives both encouragement and urgency to introduce metagenomics for evaluation as standard of care in acute care pathways (36, 37).

## References

1. Cookson W, Cox MJ, Moffatt MF. New opportunities for managing acute and chronic lung infections. Nature reviews Microbiology. 2018;16(2):111–20.

2. Charalampous T, Alcolea-Medina A, Snell LB, Williams TGS, Batra R, Alder C, et al. Evaluating the potential for respiratory metagenomics to improve treatment of secondary infection and detection of nosocomial transmission on expanded COVID-19 intensive care units. Genome Medicine. 2021;13(1):182.

3. Wilson MR, Sample HA, Zorn KC, Arevalo S, Yu G, Neuhaus J, et al. Clinical Metagenomic Sequencing for Diagnosis of Meningitis and Encephalitis. New England Journal of Medicine. 2019;380(24):2327–40.

4. Gu W, Deng X, Lee M, Sucu YD, Arevalo S, Stryke D, et al. Rapid pathogen detection by metagenomic next-generation sequencing of infected body fluids. Nature medicine. 2021;27(1):115–24.

5. Street TL, Barker L, Sanderson ND, Kavanagh J, Hoosdally S, Cole K, et al. Optimizing DNA Extraction Methods for Nanopore Sequencing of Neisseria gonorrhoeae Directly from Urine Samples. Journal of clinical microbiology. 2020;58(3).

6. Charalampous T, Kay GL, Richardson H, Aydin A, Baldan R, Jeanes C, et al. Nanopore metagenomics enables rapid clinical diagnosis of bacterial lower respiratory infection. Nature biotechnology. 2019;37(7):783–92.

7. Mu S, Hu L, Zhang Y, Liu Y, Cui X, Zou X, et al. Prospective Evaluation of a Rapid Clinical Metagenomics Test for Bacterial Pneumonia. Frontiers in cellular and infection microbiology. 2021;11:684965.

8. Blauwkamp TA, Thair S, Rosen MJ, Blair L, Lindner MS, Vilfan ID, et al. Analytical and clinical validation of a microbial cell-free DNA sequencing test for infectious disease. Nature Microbiology. 2019;4(4):663–74.

9. Baldan R, Cliff PR, Burns S, Medina A, Smith GC, Batra R, et al. Development and evaluation of a nanopore 16S rRNA gene sequencing service for same day targeted treatment of bacterial respiratory infection in the intensive care unit. Journal of Infection. 2021;83(2):167–74.

10. Justin Joseph O’grady GLK, Themoula Charalampous, Alp Aydin, Riccardo SCOTTI, inventorMethod for digesting nucleic acid in a sample. United Kingdom patent WO2021105659A1. 2109 03/06/2021.

11. Kim D, Song L, Breitwieser FP, Salzberg SL. Centrifuge: rapid and sensitive classification of metagenomic sequences. Genome research. 2016;26(12):1721–9.

12. Sichtig H, Minogue T, Yan Y, Stefan C, Hall A, Tallon L, et al. FDA-ARGOS is a database with public quality-controlled reference genomes for diagnostic use and regulatory science. Nature Communications. 2019;10(1):3313.

13. Jeck WR, Iafrate AJ, Nardi V. Nanopore Flongle Sequencing as a Rapid, Single-Specimen Clinical Test for Fusion Detection. The Journal of Molecular Diagnostics. 2021;23(5):630–6.

14. Peacock S. Health care: Bring microbial sequencing to hospitals. Nature. 2014;509(7502):557–9.

15. Armstrong GL, MacCannell DR, Taylor J, Carleton HA, Neuhaus EB, Bradbury RS, et al. Pathogen Genomics in Public Health. New England Journal of Medicine. 2019;381(26):2569–80.

16. Yamasaki K, Kawanami T, Yatera K, Fukuda K, Noguchi S, Nagata S, et al. Significance of Anaerobes and Oral Bacteria in Community-Acquired Pneumonia. PLOS ONE. 2013;8(5):e63103.

17. Mandell LA, Niederman MS. Aspiration Pneumonia. New England Journal of Medicine. 2019;380(7):651–63.

18. Mukae H, Noguchi S, Naito K, Kawanami T, Yamasaki K, Fukuda K, et al. The Importance of Obligate Anaerobes and the Streptococcus anginosus Group in Pulmonary Abscess: A Clone Library Analysis Using Bronchoalveolar Lavage Fluid. Respiration. 2016;92(2):80–9.

19. Metlay JP, Waterer GW, Long AC, Anzueto A, Brozek J, Crothers K, et al. Diagnosis and Treatment of Adults with Community-acquired Pneumonia. An Official Clinical Practice Guideline of the American Thoracic Society and Infectious Diseases Society of America. American Journal of Respiratory and Critical Care Medicine. 2019;200(7):e45–e67.

20. Guo Y, Li L, Li Z, Sun L, Wang H. Tropheryma whipplei Detection by Nanopore Sequencing in Patients With Interstitial Lung Disease. Front Microbiol. 2021;12:760696.

21. Qin S, Clausen E, Nouraie SM, Kingsley L, McMahon D, Kleerup E, et al. Tropheryma whipplei colonization in HIV-infected individuals is not associated with lung function or inflammation. PLoS One. 2018;13(10):e0205065.

22. Zhang WM, Xu L. Pulmonary parenchymal involvement caused by Tropheryma whipplei. Open medicine (Warsaw, Poland). 2021;16(1):843–6.

23. Tatsuno I, Isaka M, Matsumoto M, Hasegawa T. Prevalence of emm1 Streptococcus pyogenes having a novel type of genomic composition. Microbiology and immunology. 2019;63(10):413–26.

24. Agency UHS. Group A streptococcal infections: report on seasonal activity in England, 2022 to 2023. UKHSA; 2023.

25. Jain N, Lansiaux E, Reinis A. Group A streptococcal (GAS) infections amongst children in Europe: Taming the rising tide. New microbes and new infections. 2023;51:101071.

26. Street TL, Sanderson ND, Kolenda C, Kavanagh J, Pickford H, Hoosdally S, et al. Clinical Metagenomic Sequencing for Species Identification and Antimicrobial Resistance Prediction in Orthopedic Device Infection. Journal of clinical microbiology. 2022;60(4):e0215621.

27. Chiu CY, Miller SA. Clinical metagenomics. Nature Reviews Genetics. 2019;20(6):341–55.

28. Alcolea-Medina A, Charalampous T, Snell LB, Aydin A, Alder C, Maloney G, et al. Novel, Rapid Metagenomic Method to Detect Emerging Viral Pathogens Applied to Human Monkeypox Infections. 2022.

29. Edgeworth J. Respiratory metagenomics: route to routine service.. Current Opinion in Infectious Diseases (in press). 2023

30. Govender KN, Street TL, Sanderson ND, Eyre DW. Metagenomic Sequencing as a Pathogen-Agnostic Clinical Diagnostic Tool for Infectious Diseases: a Systematic Review and Meta-analysis of Diagnostic Test Accuracy Studies. Journal of clinical microbiology. 2021;59(9):e0291620.

31. CDC. COVID-19: U.S. Impact on Antimicrobial Resistance, Special Report 2022.COVID-19 &Antimicrobial Resistance; 2022.

32. Murray CJ, Ikuta KS, Sharara F, Swetschinski L, Aguilar GR, Gray A, et al. Global burden of bacterial antimicrobial resistance in 2019: a systematic analysis. The Lancet. 2022;399(10325):629–55.

33. Laxminarayan R. The overlooked pandemic of antimicrobial resistance. The Lancet. 2022;399(10325):606–7.

34. Sulayyim HJA, Ismail R, Hamid AA, Ghafar NA. Antibiotic Resistance during COVID-19: A Systematic Review. International journal of environmental research and public health. 2022;19(19).

35. Langford BJ, Soucy JR, Leung V, So M, Kwan ATH, Portnoff JS, et al. Antibiotic resistance associated with the COVID-19 pandemic: a systematic review and meta-analysis. Clinical microbiology and infection : the official publication of the European Society of Clinical Microbiology and Infectious Diseases. 2022.

36. Topol E. Preparing the healthcare workforce to deliver the digital future. 2019;. 2019.

37. Topol E. Ground Truths [Internet]2022. Available from: https://erictopol.substack.com/p/a-culture-of-blood-cultures.

