## Supplementary material for "Routine respiratory metagenomics service for intensive care unit patients"

*These authors contributed equally

^Senior author

### Supplementary methods

#### Hospital governance processes for pilot service provision

A new service evaluation proposal for Respiratory metagenomics (RMg) testing for intensive care unit & ECMO patients) was submitted to the critical care governance committee in September 2021 under the NHS Quality Improvement and Patients Safety (QIPS) initiative. It was submitted by the ICU clinical lead, microbiology diagnostic laboratory and infectious diseases consultant. It included service objectives, supporting published data (1, 2) and experience providing a same-day 16S rRNA sequencing service for respiratory infections during the 2019/2020 winter season (3). It included a Standard Operating Procedure (SOP), data from a pre-pilot testing phase when surplus routine samples were analysed by RMg under research ethics framework and results compared with routine culture and other microbiology tests. The application included description of the end-to-end testing pathway (Figure 1C), result reporting methods, oversight processes including review by a group meeting bi-weekly including intensivists and internal and external clinical microbiologists, and with criteria for halting the service in response to incidents or service failings. Seminars on respiratory metagenomics were given at infectious diseases and intensive care educational meetings prior to starting and questionnaires were conducted for 10 representative scientists and clinicians from the clinical laboratory and the intensive care to understand their views on unmet needs that RMg could meet, their understanding of the methodology and their views on interpretation and potential utility. The service was agreed and commenced on November 22^nd^ 2021. Follow up meetings were held after the first 4 weeks before recommencing in January 4^th^ 2022 and continuing through to 25^th^ March 2022.

#### Recommencing pilot service provision during the 2022 winter season

Data from the first winter season service was reported back to the ICU governance group. Absence of severe seasonal CA-LRTI cases was noted due to provision during the unusual setting of the COVID-19 pandemic. They were an important target group identified in the QUIPS submission. The service was therefore restarted on 3^rd^ October 2022 and provided for testing all lower respiratory tract (LRT) samples taken from the ICU admitting severe CA-LRTI patients and that provided ECMO support. Service processes were the same as the previous season but without the bi-weekly oversight group. Samples were multiplexed on MinION flowcells.

#### Microbiological culture

Microbiological culture for all respiratory samples was performed in an ISO15189-accredited laboratory following standard operating procedures (4). Tracheal aspirates were streaked directly (10 μl of sample) onto blood agar and chocolate agar plates without any prior centrifugation. BALs and NDLs were initially spun down at 1200g for 10 min, with supernatant decanted and sample sediment was retained (~500 μl of sample). Residual sample was re-suspended by vortexing (10 sec), and 10 μl of sample streaked onto blood agar, chocolate agar and fastidious anaerobic agar (FAA). All plates were incubated at 37 °C in an aerobic or an anaerobic environment for 48 h for initial examination. For the detection of *Candida* spp. and *Aspergillus* spp. Sabouraud (SAB) plates were set up and incubated for 5 days at 37 °C in aerobic conditions. MALDI-TOF (Bruker) was used to confirm bacterial colonies and microscopy for *Aspergillus* spp. Sample reported ‘normal respiratory flora (NRF)’ where no pathogenic organism was reported or as ‘no growth (NG)’ when no organisms were observed after 48 h of incubation were considered as culture-negative samples in this study. Antibiotic susceptibility for any detected pathogens was performed using agar diffusion, following guidelines set up by the European Committee on Antimicrobial Susceptibility Testing (EUCAST) methodology (5).

#### Routine detection of viral and atypical pathogens

For routine detection of SARS-CoV-2, reverse transcriptase (RT) PCR using the Highplex 24 system (AusDiaganostics Pty Ltd.),was performed by the clinical laboratory according to the manufacturer’s instructions (SARS-CoV-2, Influenza and RSV 8-well, Catalogue number: 20081, version: 08). This assay utilises 200 μl of decanted supernatant from respiratory clinical samples targets the Orf1ab and Orf8 of SARS-CoV-2. For the detection of respiratory viruses and atypical pathogens such as *Legionella pneumophila*, multiplex RT-PCR was performed by the clinical laboratory upon request.

#### Routine testing of 16S bacterial rRNA gene sequencing

Samples requested for 16S bacterial rRNA gene sequencing were referred to the Great Osmond Street Hospital (GOSH). Briefly, extracted bacterial DNA was subjected to PCR amplification of the 16S rRNA bacterial gene followed by sequencing of the PCR product using an Illumina platform.

#### Galactomannan

For galactomannan (GM) requests the clinical laboratory referred samples to the Mycology Reference Laboratory National Infection Services, UKHSA at Southmead Hospital, Bristol where the Platelia *Aspergillus* Antigen kit (BIO-RAD –62794) is used. Sera and BALs were subjected for GM detection following manufacturer’s instructions.

#### RMg Sequencing workflow

RMg workflow consisted of three aspects: host DNA depletion, microbial DNA extraction and sequencing was performed based on previously published (1, 2) and patented methods (6). In total 128 samples were processed for RMg sequencing which included 111 bronchoalveolar lavages (BAL), 3 tracheal aspirates, 8 non-direct bronchoalveolar lavages (NDL) and 6 pleural fluids (PF). Sample anonymization was done prior to RMg processing (winter 2021-2022 samples = P1-P128 and winter 2022-2023 samples= CS016, CS026, CS034, CS045, and CS053).

Initially, mucoid respiratory samples only, were sputasol-treated (SR0233 - Oxoid) in a 1:1 ratio for 15 min at 37°C to homogenise samples. For non-mucoid samples, 1 ml of sample was used for RMg sequencing.

At this step a competitive spiked-in internal control (IC) was introduced in each sample at the beginning of each run along with a no-template-control (NTC) and a positive control (PC) with all controls processed through the full pipeline. For NTC, sample was replaced with nuclease free water (NFW) and introduced to monitor barcode cross-talk and laboratory and/or reagent contamination. For the IC, 10^4^ cfu/ml of *Jonesia denitrificans* (JD)) (NCTC 10816) was introduced into each patient sample – this allowed identifications of individual sample failures. Spiked quantity of JD was predetermined after serial dilutions of the organism were spiked in culture-positive and culture-negative samples. Chosen quantity allowed identification of no-growth samples without affecting detection of potential pathogens in positive samples (data not shown). For the PC, 10^4^ cfu/ml of JD was introduced in a PBS control which acted as a positive control to monitor whole run failures. An external positive control (EPC) was also introduced when a new batch of reagents was used and was processed in parallel with patient samples. For the EPC 10^6^ cfu/ml of environmental Gram-positive (*Aneuribacillus aneurinilyticous*: ATCC 12856) and Gram-negative bacteria (*Listonella pellagium*: NCTC 11316) and yeast (*Zygosaccharomyces rouxi*: NCPF 3879) were introduced into a PBS control. The EPC was used to ensure efficiency of new reagents.

Following sputasol treatment, samples were centrifuged at 12,000xg for 5 min and supernatant was then carefully decanted leaving ~50 µl and pellet for host depletion step. For host depletion, 200 µl of PBS and 200 µl of HL-SAN buffer (5M of NACL and 100mM of MgCl_2_) were added along with 40 µl of saponin (working stock of 1% saponin (Sigma – 47036-50G-F)) and 10 µl of HL-SAN DNase (Articzymes – 70910-202). Samples were incubated on a thermomixer at 37°C shaking at 1000 rpm for 10 min to induce host cell lysis and digest of newly-released host DNA. Samples were then washed with 800 ml of PBS and centrifuged (12,000 xg for 3min) to pellet microbial organisms.

For microbial cell lysis pellet was re-suspended in lysis buffer (800 µl; Roche UK) followed by bead-beating (Lysis Matrix E beads (MP Biomedicals) and 3 min at 50 o/s on Human Tissue Lyser, Qiagen) to release microbial DNA. Centrifugation (1 min at 18,000xg) and removal of ~200 µl supernatant were then followed. Next, supernatant was treated with proteinase K (20 µl; Qiagen UK) for 5 min at 65°C shaking at 1000 rpm; on a thermomixer to digest residual proteins followed by DNA extraction using the Fast Pathogen 200 protocol on a MagNA Pure 24 System (Roche UK).

Prior to library preparation samples and controls were subjected to individual 1.2X AMPure XP bead wash and eluted in 15 µl of NFW. For each daily run (which included ≤3 samples plus controls) library preparation was performed using the Rapid PCR Barcoding Kit (Oxford Nnaopore Technologies (ONT)) as previously described (1, 2) with following alterations; for the PCR reaction Taq DNA polymerase (PrimeSTAR® GXL DNA Polymerase TaKaRa) with double reaction volumes was used with the following conditions; initial denaturation at 98 °C for 2 min, cycling conditions for 35 cycles were denaturation at 98 °C for 15sec, annealing at 56 °C for 15sec, extension at 68 °C for 45 sec °C and final extension at 68 for 4 min. Following PCR reaction, samples and controls were subjected to individual 0.6X AMPure XP bead wash and eluted in 14 µl of buffer recommended by the manufacturer.

Finally, samples and controls were prepared for singleplex sequencing using nanopore flongles (ONT) with sequencing performed on the GridION platform (ONT). Samples processed during the 2022-2023 winter season were multiplexed (≤ 4 samples plus controls) and sequenced on a MinION flowcell on the GridION platform. Raw sequence data were acquired using the ONT MinKNOW software (version 21.05.12) with live basecalling by ONT Guppy (version 5.0.16+b9fcd7b) using the barcode-at-both-ends parameter to ensure high quality barcode assortment.

Sequencing was run for 24 hours with the first 30 min data used for pathogen identification and 2 hours data used for AMR gene detection using an in-house developed bioinformatics pipeline (<https://github.com/GSTT-CIDR/RespiratoryCMg>). Prior to analysis, human reads were discarded by alignment with genome reference (GCA_000001405.15, assembly GRCh38.p13 version) and non-human reads were exported and used for pathogen identification and AMR gene detection (described below). See Figure 1 for a schematic workflow of the RMg method.

Library preparation PCR products and 0.6X bead wash eluates were assessed by DNA quantification using the high sensitivity dsDNA assay kit (Thermo Fisher) on the Qubit 3.0 Fluorometer (Thermo Fisher) prior to nanopore sequencing.

#### Analysis of metagenomic data

##### Determining parameters for microbial classification

Microbial reads were classified against a custom microbial database using Centrifuge ((7)v1.0.4) with default parameters. Reads mapping to a single organism were binned into species-level taxonomic groups for abundance estimation. Reads mapping to multiple organisms were aligned to their respective genome assemblies with minimap2 ((8), v2.18) with the read assigned to the taxonomic group with the best alignment based on BLAST identity score.

Classification score and abundance thresholds used in this study were determined using the dataset published by Charalampous *et al*. 2021 (3) as the training set. A series of abundance thresholds (≥ 0.1%, 1%, 2%, 5%, 10%) and classification scores provided by Centrifuge (≥2000, 8000, 16000) were tested to determine the best thresholds for bacterial detection to use in order to ensure the best performance for detection of true-positive findings whilst reducing identification of false positive and false negative findings.

For fungal and yeast detection a more sensitive threshold was required. A threshold of ≥5 for *Aspergillus* and *Candida* spp. classified reads was used to determine any sample positive for yeast or fungal organisms. This was based on previous knowledge that the clinical microbiology laboratory tests utilise more sensitive detection limits for fungal and yeast organisms – if a single *Aspergillus* colony is identified on the cultivated plate then sample is considered positive for *Aspergillus*.­ This differs from the more stringent detection of microbial culture used for bacterial organisms (~10^2^ bacterial cfu/ml – i.e. one colony identified on a plate previously streaked with a swab taken from an undiluted LRT sample). Applying a less-stringent threshold provided the highest sensitivity while avoiding false positive detections.

The parameters chosen were then tested on a number of samples (n=18) and quality controls were processed in a pre-pilot phase prior to the pilot study and results were compared to routine culture results (Table S6). Sensitivity and specificity was calculated on a per sample basis (14) using the Clopper–Pearson exact method (<https://www.medcalc.org/calc/diagnostic_test.php>) (Table S7).

Based on these findings in order to not compromise sensitivity and specificity of the method the chosen single-read centrifuge classification score was >7999 with reads with <8000 centrifuge score removed from further analysis. Similarly, bacterial organisms represent ≥1% of total passed microbial reads. Fungi and yeast were reported if ≥ 5 passed reads with a >7999 centrifuge score were identified.

##### Database for microbial classification

The FDA-ARGOS microbial reference database (9), was used to create the Centrifuge index used for microbial identification. The FDA-ARGOS genomic database utilises numerous quality-control metrics based on the regulatory-grade reference genome criteria that a successful genome entry would require. For example, required coverage for genome assembly is 95% with 20X depth at every position across the assembly (9). A list of clinically-relevant organisms was compiled based on previous microbiology results reported in the past 5 years. Based on this list the database was further curated with organisms either added or removed from the database (<https://github.com/GSTT-CIDR/RespiratoryCMg>). Genome assembly data retrieved from Refseq database (9, 10) and Dustmasker (version 2.10.1) was used to mask low complexity regions. The “centrifuge_build” tool from Centrifuge was used on default parameters to create the index. The database consists of 673 entries which includes 624 bacteria, 3 DNA viruses, 45 fungi and yeast and 1 protozoa ((<https://github.com/GSTT-CIDR/RespiratoryCMg>).

##### Misclassification rate

The misclassification of reads to closely-related species is a common bioinformatics problem in metagenomics. To address this, a selection of commonly-found organisms in LRT samples (*Enterobacter cloacae, Enterococcus faecalis, Enterococcus faecium, Escherichia coli, Klebsiella aerogenes, Klebsiella oxytoca, Klebsiella pneumoniae, Klebsiella variicola, Staphylococcus aureus, Staphylococcus epidermidis, Streptococcus mitis, Streptococcus oralis, Streptococcus pneumoniae*) were chosen and data from isolate whole genome sequencing (WGS) were retrieved from NCBI (Table S8). Data was subsampled to 200mbases of sequencing reads and then processed using the in-house metagenomics pipeline to assess the True positive (TP) and false positive (FP) rate.

A truth set was derived from mapping reads to reference genomes using minimap2. Reads were given a classification category based on the concordance (TP, True Negative (TN)) or discordance (FP, and False Negative (FN)) between the in-house metagenomics pipeline and minimap2 (Table S7). The percentage of TP classified reads was ≥ 93% for *E. cloacae* (94.6%)*, E. faecalis* (95.1%)*, E. faecium* (94.5%)*, E. coli* (94.0%)*, K. aerogenes* (94.5%)*, K. pneumoniae* (93.7%)*, K. variicola* (96.1%)*, S. aureus* (96.6%)*, S. epidermidis* (94.2%) and *S. pneumoniae* (99.0%), whilst a lower percentage of TP classified reads of ≥70% was achieved for *S. mitis* (70.0%), *S. oralis* (79.9%) and *K. oxytoca* (77.9%). For *S. mitis, S. oralis* the percentage of TP reads at the genus level increases to 90.2% and 98.2% respectively, indicating that the majority of misclassified reads happen at an intra-genus level. *K. oxytoca* contained a high number of reads classified as TN (12.1%) indicating a large level of contamination in the dataset.

Therefore in order to address the misclassification rate observed, the rate of TP classified reads were used in the algorithm development for realignment of misclassified reads within our metagenomic datasets. Reads were first binned into genus-level groups and the abundance of each species calculated within these bins. The most abundant species for each genus bin were identified, and assigned the reads for that genus if its genus-level abundance was higher above the threshold determined above. If the genus-level abundance for the top species fell below this threshold, then the next most abundance specie(s) were added until the threshold was met. Reads within the bin were then distributed proportionally across these organisms.

Reads were determined to be TP if they mapped to the correct species using the in-house metagenomics pipeline and mapped in minimap2. TN reads were identified if they were classified as a different species with the in-house metagenomics pipeline and not mapped in minimap2 and reads were determined as FN if they mapped to a non-target species in with the in-house metagenomics pipeline but mapped in minimap2. Finally, reads were determined to be FP if they mapped to target species with the in-house metagenomics pipeline but not mapped in minimap2)**.**

##### Phenotypic prediction based on detected resistance genes

Phenotypic prediction was performed based on the detection of resistance genes using 2 hours of sequencing data. For the detection of antibacterial resistance genes Abricate (11) and Scagaire (12) were used both with default parameters. Briefly, basecalled and demultiplexed FASTQ files were converted into FASTA files and were analysed using Abricate, to detect resistance genes against the Comprehensive Antibiotic Resistance Database (CARD) (13). Next, based on the Abricate output, Scagaire was used to predict and include only clinically-relevant genes based on the pathogen identified by RMg. Scagaire uses a bundled database consisted of the 40 most common-sequenced bacterial species present in the RefSeq database and is designed to only report clinically-relevant resistance genes. Clinically-relevant gene alignments were reported only if >1 gene alignment was detected with <90% gene coverage to ensure exclusion of any possible bioinformatics errors.

This analysis was only carried out to determine presence or absence of acquired genotypic determinants conferring resistance to antibiotics used on the ICU. These included (i) genes conferring resistance to beta-lactams in Enterobacterales (ii) presence of *mecA* genes in *S. aureus*-positive samples which would confer resistance to Methicillin and (iii) presence of *van* gene clusters in *E. faecium*-positive samples which would confer resistance to Vancomycin (Table S9).

##### Reporting of detected organisms and resistance gene

The pipeline generated sequencing reports at three time intervals (30min, 2 hrs and 16 hrs of sequencing) which were interpreted following the Standard Operation Procedure (SOP) for reporting metagenomics results previously formulated by clinicians and scientists.

The sequencing report was divided into three sections, (i) sample identifiers, (ii) quality control measurements, (iii) microbial organisms and resistance genes detected above pre-defined thresholds and (iv)full list of classified organisms and AMR genes detected before applying pre-defined thresholds.

The reporting SOP described quality control thresholds and positivity thresholds established using pre-existing RMg data produced from our workflow. Initially, each sample was assessed based on performance, quality and contamination rate. Samples with <100 total reads and/or <10 microbial classified reads after 2hrs of sequencing were failed and repeated on the next day if residual sample was available. Samples with common contaminant reads representing ≥10% of total microbial classified reads were considered contaminated and also failed*.* Samples with ≥100 reads after 2hrs and <10% contamination rate were passed and reported accordingly based on the organisms and resistance genes detected. For samples where only the IC control organism (JD) was detected were reported as negative.

Quality controls introduced at the beginning of each sequencing run were also assessed. For the PC, *J. denitrificans* reads were recorded after every daily run during the pre-pilot and pilot phase in order to establish a cut-off demonstrating a successful run. This was used as quality control on the 30 min reports. As per latest analysis, PC control with ≥100 classified *J. denitrificans* reads and were consisted 90% of the total microbial classified reads would pass the quality control (QC) check. For the NTC, any reads identified in the NTC were considered contaminants and if were present in samples were also considered contaminant reads and were excluded. Common contaminants in our dataset mainly included environmental organisms such as: *Moraxella osloensis, Cutibacterium acnes* and *Acinetobacter johnosonii* and *E. coli.* As *E. coli* was a common contaminant in our dataset, a sample was only considered *E. coli*-positive only if *E. coli* reads were more abundant than the reads assigned to the IC. This allowed the identification of true-positive findings whilst eliminating contaminant reads.

A list of reportable organisms was compiled and followed for reporting. The list was based on previous lower respiratory tract infections studies (1, 2, 14-16) and previous findings from the archives of microbiological culture in the last 5 years collected from the clinical laboratory (Table S10). Microorganisms reported from the respiratory RMg workflow in this study were referred to as ‘respiratory pathogens’ or ‘pathogens’ and they were defined as microorganisms causing respiratory infection in ICU patients.

Detected respiratory pathogens samples processed in the pilot study were: *Aspergillus flavus*, *Bulkhoderia* spp., *Citrobacter koseri*, *Citrobacter freundii*, *Escherichia coli*, *Enterobacter cloacae* complex, *Fusobacterium nucleatum*, *Hafnia alvei*, *Klebsiella aerogenes*, *Klebsiella michiganensis,* *Klebsiella oxytoca*, *Klebsiella pneumoniae,* *Klebsiella variicola, Legionella pneumophila,* *Morganella morganii*, *Mycobacterium tuberculosis*, *Proteus mirabilis*, *Pseudomonas aeruginosa*, *Serratia marcescens*, *Stenotrophomona maltophilia*, *Staphylococcus aureus*, *Streptococcus pneumoniae and Tropheryma whipplei*.

*Reporting criteria for certain organisms were also followed based on criteria set by the clinical laboratory for microbiological cultures.* Anaerobes were only reported if above pre-defined thresholds in BALs and PFs. *Candida* spp. is often detected in LRT samples but considered a common lung coloniser and not an infectious agent (17). Hence, *Candida* spp. were reported following fungal-specific thresholds but were not defined as a ‘pathogen’ in this study. Clinical-relevance of *Corynebacterium striatum* and *Enterococcus faecium* was also unclear. Hence, *C. striatum* and *E. faecium* along with other organisms with the “if predominant” flag in Table S10 were only reported to the clinical team if they were the highest abundant organism with respect to normal flora detected within the sample.

#### Sequence-based typing of organisms for infection control purposes using sequencing data

Downstream analysis was performed for organisms identified from metagenomic sequencing that were suspected in transmission events. The analysis included reference-based alignment against a representative genome based on organism of interest, generation of consensus sequence using aligned reads, SNP distance calculation between the consensus sequences (1). For this analysis 24hrs of sequencing RMg data were used and genomes were considered related if a genetic similarity of ≥99.99% was reported <https://nanoporetech.com/accuracy>. Pathogens from patients not included in the study but were suspected to be part of an outbreak were isolated, extracted and subjected to whole genome sequencing as previously described (1) to investigate genetic similarity from pathogens identified in samples processed with RMg.

### Supplementary Results

#### Limit of Detection (LoD) of the RMg workflow

We previously reported that the analytical LoD of the RMg workflow is dependable on the human/microbial DNA levels present in the each sample (LoD=range of 10^3^-10^5^ c.f.u/ml for respiratory samples) (2). However, we sought to ensure that the introduction of the IC organism did not affect the analytical LoD of the method. Therefore, to determine the analytical LoD of the method, an uninfected ‘upper respiratory tract flora’ (URTF) BAL sample was used. Serial tenfold dilutions of Gram negative (*K. pneumoniae*), Gram positive (*S. aureus*) and a yeast (*Candida albicans*) were introduced to the aliquoted triplicates of the BAL sample. The IC organism was also introduced in pre-determined cell-quantities (10^4^ c.f.u/ml) in each replicate. The LoD (≥2/3 positive replicates) for both bacterial organisms (*K. pneumoniae* and *S. aureus*) and the yeast (*C.* albicans) in the BAL sample was determined to be 1000 (10^3^) cells after applying pre-defined thresholds. Hence, the analytical LoD of the method was determined to be 1000 c.f.u/ml for bacteria and yeast (Table S11).

#### Pre-pilot runs

In total 18 samples spiked with the IC organism were depleted and sequenced along with QC as previously described. The pre-pilot was carried out to ensure all aspects of the workflow which includes host depletion, library preparation, nanopore sequencing and data analysis were performing well (see figure 1).

In this sample set 13/18 samples were concordant with the culture including 11 culture-positive samples and 2/14 reported as culture-negative or with not-clinically significant organisms by culture (Table S6A). The remaining samples (n=5) were deemed discordant. These included two samples were RMg missed the culture-reported organism (PR9 and PR12) and two culture-negative samples where metagenomics identified additional organisms – Vancomycin-resistant Enterococcus faecium (VRE) in PR6 and Streptococcus pneumoniae in PR14. Both additional findings were confirmed by additional testing. The last discordant sample did not produce enough reads during RMg sequencing so it was deemed as a failure.

The pre-pilot dataset was used as the testing dataset to test positivity thresholds and quality-control rules. Also, analysis of the quality controls revealed that *E. coli* contamination was common in the pre-pilot sample set, hence the reporting rule for *E. coli* was used as described previously – *E. coli* would only reported if is present at a higher abundance from the internal control organism in clinical samples (Table S6B). Additionally, applying QC rules, allowed identification of failed sample (PR13 had <100 total reads after 2hrs of sequencing) and failed positive control. Only one PC failed – PR-C7 had <100 total reads after 2hrs of sequencing.

#### Technical failures

In total there were 128 respiratory samples processed with the RMg workflow. However, 15/128 (12%) samples failed during QC and where excluded from the study (Table S2). The majority of the samples failed due to microbial reads being ≤10 reads after 2hrs of sequencing and hence failed QC checks and were excluded from reporting and downstream analysis (n=12/15 samples). The increased failure rate was caused by using a faulty batch of extraction kit (confirmed by the manufacturer) that resulted to a tenfold microbial loss after extraction. The batch was used during the pilot for the first two months which possibly led to the failure of 9/15 samples. Majority of the samples were also reported as ‘no organisms detected’ by microbiological testing which reflected that microbial loss caused from faulty extraction batch could have led to the samples below the LoD of the RMg workflow.

### Supplementary figure footnotes

#### Supplementary footnote to patients’ timelines for Figure 2.

**A)	ICU acquired *L. pneumophila* ST1326 pneumonia. Unexpected bacteria prompting antibiotic escalation, infection control and public health interventions.**

55-60 year old woman post aortic and mitral valve replacement. Extubated post operation but increasing secretions and heart failure requiring re-intubation. Sepsis was considered a contributory factor, with empiric vancomycin and gentamicin started due to penicillin allergy. Two days later temperature had increased to 38.5, inotropes started and CT chest showed new consolidation. RMg identified *L. pneumophila* at 30 minutes sequencing. The patient had been started on levofloxacin prior to communication due to continued deterioration. There was sufficient sequence for MLST at 8 hours to identify a non-serogroup strain based on detecting 5 of 7 MLST gene (flaA[3], PilE [10], asd [-], Mip [28], mompS [-], proA [9], neuA [207]). Sample was confirmed by local PCR and culture, but not urinary legionella antigen testing, with typing confirmed on a cultured colony from the respiratory sample as ST1326 by reference laboratory after 7 days. Infection control was informed on day of RMg result leading to declaration of a serious incident and urgent water sampling. *L. pneumophila* ST1326 found in surgical ICU tap water sent to the public health reference laboratory with highest concentration in water from tap adjacent to patient bed space [21]. Water filters were fitted onto all ICU water taps pending definitive resolution.

**B)	*P. aeruginosa* VAP. New bacterial pathogen in patient with severe COVID-19 pneumonitis prompting antibiotic escalation**

40-45 year old man who was SARS-CoV-2 unvaccinated with no past medical history or risk factors for severe COVID-19. Referred for ECMO few days after ICU admission with severe COVID-19 pneumonitis and a right hydro-pneumothorax and 2 intercostal drains. Admission BAL grew only scanty *S. marcescens* so linezolid was stopped and meropenem continued. A repeat BAL taken few days after ICU admission reported scanty S. maltophilia on Day 6 and with CRP increasing meropenem was changed to septrin (the *S. marcescens* was also reported as septrin susceptible). Tracheostomy performed and further intercostal drain inserted and patient made little progress with drains intermittently blocked. Repeat BAL for culture again identified the *S. maltophilia* and *S. marcescens*. RMg was performed two weeks post-admission to inform whether to either prolong the septrin course or escalate treatment. Identification of *P. aeruginosa* by RMg informed change to meropenem and gentamicin. *P. aeruginosa* was not grown from the contemporaneous sample submitted for culture that day but was grown from a follow up BAL. The patient made steady progress over a prolonged ICU stay leading to successful discharge. Escalation to appropriate antibiotics informed by same day RMg identification of *P. aeruginosa* was recorded as a significant benefit by the MDT at the time.

**C)	ICU acquired vancomycin resistant *E. faecium.* Unexpected AMR-bacteria with patient & infection control impact.**

80-85 year old woman with emergency abdominal aortic aneurysm. Long term smoker, transferred to ICU for ventilation and renal replacement therapy required post-operation. Commenced on co-amoxiclav escalated to tazocin after CT chest showed bronchiectasis and consolidation. Inflammatory markers increased on tazocin so escalated to meropenem despite culture and RMg-A only showing *C. albicans*. Repeat testing (RMg-B) showing only *C. albicans* informed stopping meropenem, but patient failed to progress with CRP increasing. Chest was again considered the most likely focus so RMg-C was performed to guide therapy. Detection of VRE prompted monotherapy with linezolid associated with CRP fall from 262 to 60 and clinical improvement. The significance of VRE as a cause of VAP was debated but for this patient the RMg result was considered to have a positive impact based on the clinical response to linezolid. Infection control were informed of the result and contact precautions were instituted along with repeat VRE rectal screening that was positive.

**D)	Unexpected disseminated reactivation of HSV-2**

65-70 year old man was admitted for elective thoraco-abdominal aorta replacement. He had cardiac arrest in theatre and massive blood transfusion. He had no history of immunosuppression. He was initially extubated but then re-intubated in the surgical recovery unit around day 5 and commenced empirically on piperacillin-tazobactam for presumed HA-LRTI. A BAL taken at intubation grew *C. albicans*. He initially improved but then deteriorated ~two-weeks post-surgery with new pyrexia (40^o^C) and was transferred to the ICU where a repeat BAL was taken and he was empirically started on meropenem and anidulafungin. The BAL grew *C. glabrata*. He did not progress and was reported as being obtunded when sedation was lightened and had a new maculopapular rash on abdomen and thighs. He also had persistent temperature with increasing inflammatory markers with bilirubin raised at 113 and ALT at 106. A BAL was sent for RMg as part of a septic screen to investigate an infectious cause, although HA-LRTI was not considered the most likely explanation. When HSV-2 was identified (99% of reads), a decision was made to start high dose acyclovir and a plasma HSV PCR was requested. Dermatology review suspected a drug rash and meropenem was stopped. The rash was reviewed by virology and a vesicle identified which was swabbed along with the surgical wound. Plasma, wound and vesicle swabs were all HSV-2 PCR positive. The patient gradually improved over the following days and CRP falling from 335 to 172.

**E)	Influenza with secondary *S. pyogenes* infection not detected by culture**

65-70 year old man with no past medical history presented with history of cough and sore throat. Intubated on arrival in emergency department (ED) and transferred to referral hospital ICU but failed conventional mechanical ventilation that day so transferred to St Thomas’ for ECMO. Upon arrival he was shocked but passing urine and had high inflammatory markers. An influenza PCR was positive. He was stabilised, treated with zanamavir, meropenem, clarithromycin and anidulafungin and a BAL was performed at 08:50 for RMg and culture. The RMg report was communicated at 17:20 with 13 reads of *S. pyogenes*. It was reported to intensivists who started linezolid that night and stopped clarithromycin and anidulafungin. Infection control and public health was told of an invasive *S. pyogenes* infection that day. The BAL and blood cultures taken from admitting hospital did not grow *S. pyogenes*. There were insufficient reads after 16 hour sequencing to perform genomic typing.

**F)	Influenza with secondary PVL-MRSA and *S. pyogenes* infection, the latter not detected by culture**

50-55 year old man with no past medical history presented with breathlessness, cough and chest pain. Intubated on arrival in ED and transferred to referral hospital ICU but failed conventional mechanical ventilation that day so transferred to St Thomas’ for VVA ECMO. He arrived in septic shock and multi-organ failure with rising INR and low platelets. An influenza PCR was positive. He was stabilised and treated with zanamavir, meropenem, linezolid, clarithromycin and anidulafungin. A BAL performed on next day in the morning was sent for RMg and culture. The RMg report was communicated in the afternoon identifying *S. aureus* and *S. pyogenes*. There were 22 reads of mecA that informed communication of an MRSA and 3 reads of luk F/S which informed communication of the MRSA being PVL positive. These results were communicated to hospital infection control team that day. ermC and FusC were noted but not communicated to the clinical team in line with the SOP. The patient was commenced on intravenous immunoglobin that day with further modifications to therapy over the following days. The following morning after 16 hour sequencing the MRSA was identified as ST152 and the full results were communicated to public health. Final culture result was communicated on day 5 as MRSA resistant to clindamycin and fucidic acid. The MRSA was sent to the reference laboratory for PVL typing that confirmed on day 15. 8 weeks later UKHSA reported an increase in invasive *S. pyogenes* infections. *S. pyogenes* sequence from the 16 hour report were therefore reviewed and it was identified as an emm1-M1uk clone, which has been associated with severe disease in this outbreak.

**G)	Influenza with secondary PVL MSSA and S. dysgalactiae infection the latter not detected by culture**

50-55 year old man with Crohn’s disease on azathioprine was unwell while on holiday and transferred by ambulance. On arrival in the UK he was initially transferred to an ICU for conventional mechanical ventilation but when this failed he was referred for VVA ECMO. On arrival he was shocked with multi-organ failure. An influenza PCR was positive. He was treated with Zanamavir, Meropenem, Linezolid, Clarithromycin and Anidulafungin. A BAL could not be taken on the first full day of admission due to continued haemodynamic instability but was performed the following day in the morning and sent for RMg and culture. The RMg report was interpreted as containing *S. dysgalactiae* and *S. aureus*. No *mecA* genes were detected but there were of luk F/S gene alignments were detected. PVL MSSA and *S. dysgalactiae* and were communicated in the afternoon. 119 ermC reads were noted but not reported. Meropenem and Clarithromycin were stopped the following morning and the patient was started on Flucloxacillin in addition to continuing the linezolid and anidulafungin. The PVL–MSSA was communicated to infection control and public health. Culture of the sample reported only MSSA resistant to clarithromycin. Following identification of *S. pyogenes* in case G as an emm1-M1uk clone, the S. dysgalactiae reads from the 16 hour report were reviewed and identified as subspecies equisimilis (STG62647A), which has been identified as associated with severe disease.

**H)	Influenza with secondary invasive aspergillosis prompting urgent treatment**

30-35 year old woman with a history of asthma presented to local hospital with history of cough and green/rusty sputum and influenza PCR-positive on admission. She was treated with co-amoxiclav and clarithromycin but progressed to mechanical ventilation which failed after few days prompting referral for ECMO. Culture of respiratory sample at referral hospital was negative. On arrival for ECMO, she was haemodynamically stable and commenced on piperacillin-tazobactam and zanamavir. Secondary infection was not clinically suspected. A BAL was taken on the morning of the first full day on ICU. RMg results were communicated in the afternoon as containing *Aspergillus fumigatus* based on presence of 9852 reads after 30 minutes sequencing. She was commenced on ambisome. And piperacillin-tazobactam was stopped the following morning. The following morning full *Aspergillus* genome was assembled with 82% coverage at 10x depth. Two mutations in Cyp51A (R279T and L272I) not associated with azole resistance were detected (https://sbi.hki-jena.de/FunResDb/) however this was not communicated to the clinical team. On the 5th day the culture report was scanty *Aspergillus flavus* and scanty *P. aeruginosa*. The significance of *P. aeruginosa* was unclear and the patient had continued to improve with de-cannulation targeted for few days later but piperacillin-tazobactam was started empirically. Voriconazole was commenced and the ambisome stopped when voriconazole levels were in the therapeutic range and the patient had been decannulated from ECMO. The reference laboratory confirmed *A. fumigatus* susceptible to Ambisome and Voriconazole.

### Footnote to clinical metadata file S2 columns U-Z, AA and AB

**Column Q:** CA-LRTI was designated when RMg samples were taken as part of investigation for the community-onset infection episode (Mostly day 1-3 of ICU admission but up to day 5). HA-LRTI (new) was designated when RMg was performed at the start of a new infection episode or HA-LRTI (D), when RMg was performed during an episode of HA-LRTI to investigate suspected failure of therapy or to inform whether to stop or change antimicrobials. The “unclear or other” focus was where CA-LRTI or HA-LRTI were not the primary clinically suspected focus but was part of the differential and when results from RMg would be taken into account when deciding on treatment.

**Columns R-T** were the antimicrobials prescribed at the start of the day and those that were either started or stopped in response to the RMg result either that day or the next day. Antimicrobials stopped or started for other reasons were not included. There was usually no consensus on the reasons for commencing antifungal therapy linked with identification of *Candida spp* in respiratory samples and their prescription was only recorded where there was a majority agreement that they were significant and the RMg result was the main reason for commencing or stopping treatment.

**Column U:** Brief details from discussion at time of result communication or as recorded in clinical notes at that time. Reasons for submitting samples for RMg and prescribing decisions made were not assessed against formal criteria for diagnosing CA or HA-LRTI. Interpretation of the significance of RMg identified organisms was discussed with the infectious diseases team when the result was returned but the final decision including prescribing decisions were made by the intensivist. Their decisions made in real-time were recognised to be challenging and based on sometimes incomplete information in acutely unwell patients whose condition changed rapidly. This was particularly relevant to COVID-19 patients who were often heavily immunosuppressed and had prolonged stay on ICU. Serum CRP levels are presented because they were frequently used to assess patient trajectory during a septic episode and part of the decision on interpreting significance of microbiological results.

**Column V:** Escalate (E) and de-escalate (D) was based on starting (column S) or stopping (column T) antibiotics based on RMg results. It was recognised that some opportunities for de-escalation were not taken given this was a new test under evaluation. Any bacteria subsequently identified in a respiratory sample on ICU after de-escalation was in column Q or column W. Reassure (R) was based on there either being a plan to escalate that was prevented by the RMg result or when intensivists stated when requesting RMg testing that they wanted confidence antimicrobials were active against the organisms in the LRT, or to ensure there was no new organism when the patient was not responding as expected. No impact was recorded when RMg result were not taken into account in making decisions, either because of delay or when the result was not acted on or did not address the need.

**Column W:** Comments made after follow up or at the bi-weekly service review group that considered subsequent course of patients who had antibiotics de-escalated or where there was discrepancy between RMg and culture results. Intensivists also had opportunity to explain reasons why they or their colleagues concluded that results were either reassuring (R) or of no benefit (N). There was opportunity for external microbiologists to challenge conclusions and categories. Consensus was reached in most cases and where not an explanation was recorded. Examples of discussion on how results informed the potential utility beyond antimicrobial prescribing and for future implementation were also recorded.

### Figure S1. Performance of RMg workflow against conventional testing

Performance of RMg workflow against microbiological findings from the routine laboratory and breakdown of organisms detected by metagenomics in the 2021-2022 sample cohort.

### Table S1. Microbiological culture results from all respiratory samples from all patients admitted during the pilot study period

| Bacteria in 442 respiratory samples from the whole 172 patient cohort | Number of patients with ≥1 positive sample | Number (%) of patients with ≥1 sample having acquired β-lactam resistance (%)^1^ | Phenotypic acquired β-lactam resistance |
| --- | --- | --- | --- |
| *P. aeruginosa* | 18 | 5 (3%) | MEP (2)  PTZ, CTZ (1)  PTZ, MEP (1)  MEP, PTZ, CTZ (1) |
| *E. coli* | 16 | 9 (5%) | COAM (7);  COAM, PTZ (1)  ESBL (1) |
| *K. pneumoniae* | 16 | 6 (4%) | COAM;  COAM, PTZ (3)  ESBL(2) |
| Other *Klebsiella spp* ^2^ | 8 | 0 | 0 |
| *C. koseri* | 8 | 1 | CFX, PTZ (1) |
| *S maltophilia* | 5 | 0 | 0 |
| *H. influenza* | 5 | 2 | COAM (2) |
| *P. mirabilis* | 1 | 1 | COAM (1) |
| Other *Enterobacterales*^3^ | 26 | 3 | ESBL (1)  COAM, PTZ (1)  CTZ, PTZ (1) |
| Other GNB^4^ | 4 | N/A | N/A |
| Patients with Gram negative bacteria (GNB) in any sample | 107 (62%) | 25 (23%) | COAM (15:9%): PTZ (9:5%) |
| *S. aureus* (MRSA) | 20 (0) | N/A | N/A |
| *E. faecium* (VRE) | 10 (3) | N/A | N/A |
| *E. faecalis* | 6 | N/A | N/A |
| Other Gram positive | 5 | N/A | N/A |
| Patients with Gram positive bacteria (GPB) in any sample | 38 (23%) | N/A | N/A |

88 (21%) samples were taken during the first 48 hours of admission and 334 (79%) taken >48 hours after admission. Other phenotypically reported acquired resistance ^1^ Quinolone resistance in 6 patients: *P. aeruginosa* (3), *E. coli* (1), *M. morganii* (1) C. koseri (1). Aminoglycoside resistance in 1 patient (*E. col*i) ^2^ *K. variicola* (5) *K. oxytoca* (3) ^3^ *Serratia spp.* (9) *Enterobacter spp.* (7); *M. morganii* (2); *H. alvei* (2) ^4^ 2 *Acinetobacter spp*. (2) *Achromobacter spp.* (1) *Elizabethkingae spp.* (1). Meropenem (MEP), piperacillin-tazobactam (PTZ), ceftazidime (CTZ), co-amoxiclav (COAM), CFX (cefuroxime), Extended-spectrum beta-lactamases ((ESBLs) (2 bla*_SHV_* and 2 bla*_CXT-M_* ))

### Table S2. Pilot samples failed to pass quality checks

| Sample ID | Total No reads | Human reads (2hrs) | Total Microbial Reads (2hrs) | Microbial classified (2hrs) | Reported Organisms /IC above threshold (2hrs) | Classified reads (2hs) | Routine Microbiological Testing |
| --- | --- | --- | --- | --- | --- | --- | --- |
| P10 | 18309 | 18294 | 15 | 6 | *K. pneumoniae J. denitrificans* | 4 2 | *K. pneumoniae*(S) |
| P12 | 19085 | 19074 | 11 | 2 | None | 0 | *S. aureus* (S) |
| P22 | 14802 | 14777 | 25 | 10 | *J. denitrificans* | 10 | No organisms detected |
| P36 | 0 | 0 | 0 | 0 | None | 0 | *C. krusei* C. albicans |
| P37 | 0 | 0 | 0 | 0 | None | 0 | No organisms detected |
| P42 | 56 | 40 | 16 | 11 | None | 0 | No organisms detected |
| P43 | 30 | 6 | 24 | 20 | *J. denitrificans* | 17 | URTF |
| P48 | 9846 | 9787 | 59 | 9 | None | 0 | No organisms detected |
| P51 | 8401 | 8366 | 35 | 10 | None | 0 | No organisms detected |
| P52 | 35497 | 2648 | 32849 | 32159 | *J. denitrificans A. johnsonii C. acnes E. coli* | 29272 1573 404 402 | *S. marcescens* (S) |
| P59 | 579 | 576 | 3 | 3 | None | 0 | No organisms detected |
| P61 | 19959 | 635 | 19324 | 18876 | *J. denitrificans A. johnsonii C. acnes F. periodonticum E. coli* | 16068 1214 699 391 281 | No organisms detected |
| P104 | 103 | 39 | 64 | 51 | *J. denitrificans* | 16 | No organisms detected |
| P107 | 10667 | 10601 | 66 | 1 | None | 0 | No organisms detected |
| P114 | 347 | 160 | 187 | 184 | *J. denitrificans* | 176 | *K. pneumoniae* |

### Table S3. Breakdown of discordant results, confirmatory analysis on discordant findings and outcome.

| **Detections pre-confirmatory analysis** | | | | **Confirmatory Analysis** | | |
| --- | --- | --- | --- | --- | --- | --- |
| **Sample ID** | RMg Detected | Culture | Outcome | Additional Clinical Microbiology Testing | Previous culture | Outcome Post-confirmatory testing |
| **P26** | URTF  Mixed anaerobes  *Streptococcus pneumoniae* | *Candida albicans*  URTF | FP | *Streptococcus* spp. *Prevotella* spp. identified by 16S rRNA sequencing *S. pneumoniae* detected by culture subsequently | Negative | TP |
| **P40** | *Staphylococcus aureus* | *Candida albicans* | FP | *Staphylococcus aureus* 16S rRNA sequencing | *Staphylococcus aureus* | TP |
| **P50** | *Staphylococcus aureus* | *Candida albicans* | FP | *Staphylococcus aureus* 16S rRNA sequencing | *Staphylococcus aureus* | TP |
| **P65** | Mixed Anaerobes  URTF | Negative | FP | *P.micra* detected in PF by culture subsequently | None | TP |
| **P66** | *Citrobacter koseri* | Negative | FP | Not requested | *Citrobacter koseri Klebsiella pneumoniae  Enterococcus faecalis* | FP |
| **P83** | *Staphylococcus aureus Candida albicans* | *Candida albicans* | FP | 16S rRNA sequencing detected *Staphylococcus aureus* | *Candida albicans* | TP |
| **P88** | VRE *Candida albicans* | *Candida albicans* | FP | Positive rectal screening for VRE | *Candida albicans* | TP |
| **P94** | URTF  Mixed anaerobes | URTF and GPC in the gram stain | FP | Not requested | None | FP |
| **P102** | *Corynebacterium striatum Candida albicans* | *Candida albicans* | FP | 16s rRNA sequencing detected *Corynebacterium striatum* | *Candida albicans* | TP |
| **P106** | *Enterococcus faecium* | Commensals | FP | *Enterococcus faecium* (positive in-house qPCR assay) | *Staphylococcus. epidermidis* | FP |
| **P128** | URTF  Mixed anaerobes  *Candida albicans* | URTF | FP | Not requested | *Tropheryma whipplei*  *C. albicans* | FP |
| **P2** | Negative | *Staphylococcus aureus* | FN | positive qPCR assay for *Staphylococcus aureus* | Negative | FN |
| **P7** | *Candida albicans Candida dubliniensis  Klebsiella pneumoniae* (below thresholds) | *Klebsiella pneumoniae* | FN | 16S rRNA sequencing detected *Enterobacteriacaes*pp. | *Klebsiella pneumoniae Candida dubliniensis* VRE *Staphylococcus epidermidis* | FN |
| **P9** | Negative | *Klebsiella pneumoniae* | FN | 16S rRNA PCR detected *Klebsiella pneumoniae* | Negative | FN |
| **P77** | Negative | *Staphylococcus aureus* | FN | Not requested | Negative | FN |

TP= True Positive, FN= False negative, FP= False Positive, TN=True Negative, VRE= Vancomycin resistant *E. faecium*

| Table S4A. Presenting characteristics and treatment decisions in response to metagenomic results in patients with CA-LRTI and with comparison to culture. | | | | |
| --- | --- | --- | --- | --- |
| **CA-LRTI** | **METAGENOMIC RESULT** | **CULTURE RESULT** | **16S rRNA SEQUENCING** | **ANTIMICROBIAL PRESCRIBING** |
| 45-50F CAP Empyema CRP >400 | **PLEURAL FLUID (P78):** *S. constellatus (44%) P. micra (13%) S. anginosus (4%) S. intermedius (4%) F. vaginae (2%)* | *S. constellatus* (+++)  *Prevotella buccae* (+++) | Not requested | Empiric co-amoxiclav. Prevented early escalation |
| 20-25M Trauma to chest and abdomen from car accident. ECMO aspiration. | **BAL (P87):** *F. gonidiafoemans* (34%) *P. melaninogenica* (10%) | *S. aureus* (+/-) | *Fusobacterium* spp.  *Prevotella* spp. | Piperacillin-tazobactam  (Extended from 3-7 days) |
| 40-45F mixed overdose. Found unconscious and aspiration | **BAL (P23):**  *P. nigrescens (34%) P. melanogenica (23%) P. micra (5%)* | URTF | *Streptococcus spp*  *Enterococcus spp*  *Prevotella spp* | Empiric tazocin. Prevented escalation |
| 80-85M Emergency surgery for hip fracture with MI. CRP 206 to 370. ? new LRTI | **BAL (P81)**: *P. micra (47%) S. anginonsus (13%) Bacteroides uniformis (6%)* | *S. milleri*  (prolonged culture) | *Bacteroides spp.*  *Prevotella spp.*  *Pyramidobacter spp.* | Co-amoxiclav course extended from 5 to 7 days. Prevented escalation |
| 56-60M Decompensated alcoholic liver disease with empyema | **BAL (P85)**: *P. micra (77%), S. meyeri (9%): F. nucleatum (5%) F. canifelinum (2%) S. intermedius (2%) C. albicans (0.1%)* | *S. milleri* (+/-)  *C. albicans* (+/-) | *Fusubacterium nucleatum* | Empiric tazocin de-escalated to co-amoxiclav  and fluconazole added |
| 70-75M MI. Reintubated after early self-extubation. T38.4 | **BAL (P94):** *R. mucilaginosa (54%), N. mucosa (15%), S. viridans (4%), P. melaninogenica (4%), S. odontolytica (4%), A. eruginos (3%): S. sanguinis (2): S. gordonii (1.3%), P nigrescens (1%)* | URTF (+) | Not requested | Empiric co-amoxiclav. Continued. Prevented escalation |
| 40-45F Sore throat followed by CAP with empyema. PCT>100 ?*S. pyogenes* | **BAL (P65)**: *P. micra (44%) P. nigrescens (3%)*  **PLEURAL FLUID (P70):** *P. micra (81%), P. nigrescens (8%)*  *P. melaninogenica (2%), F. nucleatum (0.2%)* | BAL: No growth  PLEURAL FLUID: *P. micra (*prolonged culture) | P65: Not requested  P70: *Prevotella spp*. | Excluded S. pyogenes. De-escalated meropenem & Linezolid to  co-amoxiclav (with metronidazole 7 days) |
| 35-40M COVID-19. ECMO. PCT>100  Suspected lung abscess discharging into pleural space | **BAL (P69)**: *C. dubliniensis*  **BAL (P71):** No organisms | BAL & PLEURAL FLUID:  *C. dubliniensis* (+/-) | P69: *Corynebacterium spp.*  P71: *Corynebacterium jeikeium* | Excluded abscess. Meropenem & Linezolid de-escalated to coamoxiclav for 5 days* |

**C. dubliniensis* was considered to have contaminated the lower respiratory tract and was not treated.

### Table S4B. Presenting characteristics and treatment decisions in response to metagenomic results in patients with HA-LRTI of anaerobes and with comparison to culture

| **HA-LRTI: CLINICAL DETAILS** | **METAGENOMIC RESULT** | **CULTURE** | **16S rRNA SEQUENCING** | **ANTIBIOTICS PRE-TEST** | **ANTIMICROBIALS AND CLINICAL PROGRESS** |
| --- | --- | --- | --- | --- | --- |
| 40-45M COVID-19 pneumonitis. Day 33 ECMO. CRP 4 to 124 over 2 days CT – Lung infarct or abscess | **BAL (P19):** *F. nucleatum* (66%): *F. canefelinum (29%): C. albicans* – 1 read | *C. albicans* (+/-) | *F. nucleatum & Prevotella spp* | No for 13 days then meropenem for 2 days before RMg | Metronidazole added. Prolonged course. |
| 70-75M Day 17 post thymomectomy.  T38.4 and CRP 50 to 279. ?HAP | **BAL (P128)** :*S. oralis (37%): Streptococcus spp (16%); S constellatus (7%): S. anginosis (6%) E. corrodens (3%): Veilonnella parvula (3%) P. melanogenica (2%) F. nucleatum (1%)* | *C. albicans* (+) | Not done | No for 5 days and ceftriaxone before that | Not started day of result. Following day increased oxygen requirement and plugging. More septic. Metronidazole started then co-amoxiclav added. Extubated |
| 45-50M 9 days COVID-19 pneumonitis in hospital before ECMO. CRP 12. RMg to exclude pathogens before treating with methylprednisolone. | **BAL (P26):** *N. mucosa (17%): P. melanogenica (11%): S. pneumoniae (3%): E. corodens (2%): S. mitis* (1.7%): <10 reads multiple other anaerobes: *C. albicans* – 3 reads. | *C. albicans* (+/-); URTF (+/-) | Streptococcus spp, H. parainfluenzae & Prevotella spp | No for 5 days then co-amoxiclav started 2 days before RMg | Linezolid added prior to methylprednisolone. Good response |
| 55-60M Alcoholic liver disease. Intubated 10 days. Persistent pyrexia and no progress. | **BAL (P98):** *R. mucilaginosa (3%): S. anginosus (3%): S. oralis (0.4%):* <10 reads multiple other anaerobes. | *C. dubliniensis* (+/-); URTF (+/-) | Not done | No for 7 days then ciprofloxacin started day before | More septic on ciprofloxacin. PCT increase from 0.6 to 2.2. Changed to tazocin. Extubated 3 days later |

**C. dubliniensis* was considered to have contaminated the lower respiratory tract and was not treated.

### Table S5A. Organisms reported by RMg and routine testing from 8 CA-LRTI infections during the second pilot period

| **RMg reported** | **Number** | **Reported by routine requested tests** |
| --- | --- | --- |
| *S. pyogenes* | 2 | 0 |
| *S. aureus*^1^ | 3 | 3 |
| *S. pneumoniae* | 2 | 1 |
| *S. dysgalactiae* | 1 | 0 |
| *H. influenzae* | 1 | 1 |
| *A. fumigatus* | 1 | 1^2^ |
| *L. pneumophila* | 1 | 1 (PCR-detected) |
| Other^3^ | 7 | 5 |
| **TOTAL** | **18** | **12** |

Routine testing was culture apart from *L. pneumohila* (PCR)

^1^ Two with *SCCmec* and all with *luk F/S*. ^2^ Reported as *A. flavus* by routine laboratory but confirmed as *A. fumigatus* by reference laboratory. ^3^ *Candida spp* (3), *E. coli* (1*), S. marcescens* (1), *K. pneumoniae* (1), HSV-1 (1). The significance of these organisms remained uncertain.

### Table S5B. Organisms reported by RMg and routine testing from 16 HA-LRTI* infections during the second pilot period

| **RMg reported** | **Number** | **Reported by routine requested tests** |
| --- | --- | --- |
| *Candida spp* | 6 | 5 |
| *K. pneumoniae* | 3 | 1 |
| Herpes simplex virus ^2^ | 3 | 0 ^2^ |
| Other | 4 | 4 |
| **TOTAL ^3^** | **16** | **10** |

Routine testing was culture apart from Herpes simplex viruses (PCR)

*RMg was performed on 17 samples but only 14 had parallel samples sent for culture and are presented in this table. ^2^ Two HSV-1 and one HSV-2, the latter clinically significant (Figure 3 case D). Routine PCR testing was not requested prior to reporting RMg result although the test was available. ^3^ *E. faecium* (VRE)*, S. marcescens,* anaerobes*, A. baumanii -* all single detections.

### Table S6A. Metagenomic data^^^ from samples processed during pre-pilot phase

| Sample ID | Total No reads | Microbial reads | Human reads | Detected Organisms/IC above threshold | Organism/IC reads | Organism reported by culture | Comments |
| --- | --- | --- | --- | --- | --- | --- | --- |
| PR1 | 6629 | 5087 | 512 | *P. aeruginosa Serratia marcescens A. xylosoxidans J. denitrificans* | 3183 1453 301 75 | *P. aeruginosa  S. marcescens* | Concordant |
| PR2 | 25524 | 4181 | 20898 | *P. aeruginosa C. freundii S. maltophilia J. denitrificans* | 3130 569 326 49 | *P. aeruginosa*  *S. maltophilia* | Concordant |
| PR3 | 15634 | 14951 | 149 | *Pseudomonas aeruginosa* | 14792 | *P. aeruginosa* | Concordant |
| PR4 | 25598 | 1607 | 23434 | *Rothia mucilaginosa S. anginonsus S. viridans Candida tropicalis Jonesia denitrificans* | 324 66 52 44 1076 | *C. tropicalis* | Concordant |
| PR5 | 20507 | 7080 | 13072 | *H. influenza  E. coli A. fumigatus* | 3010 2349 4 | *E. coli  H. influenzae  A. fumigatus* | Concordant |
| PR6 | 36980 | 12991 | 3737 | *E. faecium J. denitrificans* | 6934 1858 | Commensals | Discordant – FP |
| PR7 | 32767 | 13779 | 16701 | *P.aeruginosa  S. marcescens A. xylosoxidans J. denitrificans* | 3902 3570 3856 2263 | P. aeruginosa  *S. marcescens* | Concordant |
| PR8 | 18823 | 16241 | 18 | *P.aeruginosa A. xylosoxidans*  *B. cenocepacia J. denitrificans* | 15545 212 132 26 | P. aeruginosa  K. pneumoniae* | Discordant – False Negative |
| PR9 | 17854 | 10326 | 85 | *N. mucosa*  *P. melaninogenica*  *S. viridans J. denitrificans A. fumigatus* | 6378 2715 212 10 13 | *A. fumigatus* | Concordant |
| PR10 | 20599 | 11534 | 5958 | *P. aeruginosa J. denitrificans C. albicans* | 6569 333 15 | *P. aeruginosa  C. albicans* | Concordant |
| PR11 | 29609 | 24998 | 1838 | *K. aerogenes J. denitrificans* | 24165 374 | *K. aerogenes* | Concordant |
| PR12 | 6828 | 921 | 5625 | *C. freundii P. aeruginosa J. denitrificans C. koseri S. maltophilia* | 461 212 94 85 50 | *A. fumigatus** | Discordant – False Negative |
| PR13 | 50 | 16 | 22 | *J. denitrificans E. coli* | 13 2 | *P. aeruginosa  C. albicans* | fail |
| PR14 | 11771 | 121 | 11609 | *S. pneumoniae J. denitrificans* | 122 4 | Negative | Discordant – False Positive Pneumococcal antigen positive |
| PR15 | 1086 | 244 | 819 | *J. denitrificans* | 241 | Negative | Concordant |
| PR16 | 12803 | 6868 | 4300 | *P. aeruginosa J. denitrificans* | 211 6542 | *P. aeruginosa* | Concordant |
| PR17 | 14532 | 8891 | 1082 | *N. mucosa H. parahaemolyticus* | 8409 103 | URTF | Concordant |
| PR18 | 5658 | 776 | 4838 | *P. aeruginosa J. denitrificans* | 11 762 | *P. aeruginosa* | Concordant |

^ sequencing data after 2hrs of sequencing; *Organism missed by the RMg workflow

| Table S6B: Metagenomic data* of quality controls processed during the pre-pilot phase | | | | | | |
| --- | --- | --- | --- | --- | --- | --- |
| Negative control | Total No reads | Microbial reads | Human reads | Organisms Identified | Organisms reads | *J. denitrificans* abundance (%) |
| PR-PC1 | 18814 | 17149 | 62 | *J. denitrificans* | 17018 | 99.2% |
| PR-PC2 | 16833 | 13282 | 132 | *J. denitrificans P. aeruginosa* | 12904 185 | 97.1% |
| PR-PC3 | 4908 | 3138 | 55 | *J. denitrificans E. coli* | 2886 184 | 91.9% |
| PR-PC4 | 23840 | 20926 | 52 | *J. denitrificans E. coli* | 20635 190 | 98.6% |
| PR-PC5 | 21205 | 19758 | 71 | *J. denitrificans E. coli* | 19417 228 | 98% |
| PR-PC6 | 22578 | 21085 | 139 | *J. denitrificans C. acnes* | 19875 846 | 94% |
| PR-PC7 | 439 | 392 | 1 | *J. denitrificans S. mitis* | 363 14 | 92% |
| PR-PC8 | 7 | 6 | 0 | *J. denitrificans* | 6 | 100% |
| PR-PC9 | 9919 | 8927 | 15 | *J. denitrificans* | 8873 | 99% |

*sequencing data after 2hrs of sequencing

### Table S7. Performance reported after testing different parameters on training set for pathogen identification.

Abundance thresholds (≥ 0.1%, 1%, 2%, 5% and 10%) tested against a series of classification scores provided by Centrifuge. (≥ 2000 (A), ≥ 8000 (B), ≥ 16000 (C).

**A**

|  | **0.1%** | **1%** | **2%** | **5%** | **10%** |
| --- | --- | --- | --- | --- | --- |
| **TP** | 13 | 12 | 11 | 10 | 9 |
| **FN** | 1 | 2 | 3 | 4 | 5 |
| **TN** | 1 | 2 | 2 | 2 | 2 |
| **FP** | 2 | 1 | 1 | 1 | 1 |
| **Sensitivity** | 0.93 | 0.86 | 0.79 | 0.71 | 0.64 |
| **Specificity** | 0.33 | 0.67 | 0.67 | 0.67 | 0.67 |

**B**

|  | **0.1%** | **1%** | **2%** | **5%** | **10%** |
| --- | --- | --- | --- | --- | --- |
| **TP** | 13 | 12 | 11 | 10 | 9 |
| **FN** | 1 | 2 | 3 | 4 | 5 |
| **TN** | 1 | 2 | 2 | 2 | 2 |
| **FP** | 2 | 1 | 1 | 1 | 1 |
| **Sensitivity** | 0.93 | 0.86 | 0.79 | 0.71 | 0.64 |
| **Specificity** | 0.33 | 0.67 | 0.67 | 0.67 | 0.67 |

**C**

|  | **0.1%** | **1%** | **2%** | **5%** | **10%** |
| --- | --- | --- | --- | --- | --- |
| **TP** | 13 | 12 | 11 | 10 | 9 |
| **FN** | 1 | 2 | 3 | 4 | 5 |
| **TN** | 1 | 2 | 2 | 2 | 2 |
| **FP** | 2 | 1 | 1 | 1 | 1 |
| **Sensitivity** | 0.93 | 0.86 | 0.79 | 0.71 | 0.64 |
| **Specificity** | 0.33 | 0.67 | 0.67 | 0.67 | 0.67 |

### Table S8: List of organisms tested to identify the misclassification rate observed using the in-house metagenomics bioinformatics pipeline

| **Organism** | **Classification rate (%)** | **Misclassified organisms (abundance %)** |
| --- | --- | --- |
| *Enterobacter cloacae* | 94.55 | *K. pneumoniae (2.68%), E. coli (0.89%)* |
| *Enterococcus faecalis* | 98.65 | *E. avium (0.6%), P. harei (0.39%)* |
| *Enterococcus faecium* | 94.68 | *E. durans (2.74%), E. faecalis (0.87%), E. hirae (0.69%), S. aureus (0.35%)* |
| *Escherichia coli* | 97.69 | *S. flexneri (0.95%), E. tarda (0.43%), C. fruendii (0.35%)* |
| *Haemophilus parainfluenzae* | 93.45 | *H. influenza (4.57%), H. aegyyptius (0.93%), H. parahaemolyticus (0.60%)* |
| *Klebsiella aerogenes* | 97.40 | *C. koseri (1.21%), K. pneumoniae (0.94%), K. oxytoca (0.23%)* |
| *Klebsiella oxytoca* | 88.78 | *K. pneumoniae (7.15%), R. planticola (1.85%), S. plymuthica (0.92%), E. coli (0.48%)* |
| *Klebsiella pneumoniae* | 95.61 | *K. oxytoca (1.03%), S. aureus (0.95%), R. planticola (0.68%), K. quasipneumoniae (0.35%)* |
| *Klebsiella variicola* | 96.41 | *K. pneumoniae (1.93%), K. quasipneumoniae (0.75%), K. oxytoca (0.31%)* |
| *Staphylococcus aureus* | 99.71 | *N/A* |
| *Staphylococcus epidermidis* | 95.96 | *S. hominis (2.36%), S. saccharolyticus (0.55%), S. aureus (0.46%)* |
| *Streptococcus mitis* | 72.28 | *S. pneumoniae (12.85%), S. oralis (7.90%), S. ARGOS256 (1.21%)* |
| *Streptococcus oralis* | 80.29 | *S.ARGOS256 (17.15%), S. intermedius (1.15%), S. gordonii (0.70%)* |
| *Streptococcus pneumoniae* | 99.74 | *N/A* |
| *Streptococcus pyogenes* | 97.00 | *S. dysgalactiae (2.27%), S. agalactiae (0.30%)* |

### Table S9: List of antimicrobial resistance gene reported in our study

| **Gene name** | **Organisms associated** | **Affected Antibiotics** | **Mechanism** |
| --- | --- | --- | --- |
| *mecA* | *S. aureus* | Methicillin | Plasmid |
| *vanA* | *E. faecium* | Vancomycin | Plasmid |
| bla*_TEM-1_* | Enterobacterales | Narrow spectrum to beta lactams | Plasmid |
| bla*_TEM-4_* | Enterobacterales | ESBL | Plasmid |
| bla*_SHV-11_* | Enterobacterales | Narrow spectrum to beta lactams | Plasmid |
| bla*_SHV_*_-186_ | Enterobacterales | ESBL | Plasmid |
| bla*_OXA_-1_1* | Enterobacterales | Ampicillin | Plasmid |
| *bla_CTX_* (most common is bla*_CTX-M-15_* & bla*_CTX-M-15_*) | Enterobacterales | ESBL | Plasmid |

ESBL= Extended spectrum beta-lactamases

### Table S10: List of pre-defined organisms^*^ and reference for each organism

| **Bacteria** | **Report** | **Reference** |
| --- | --- | --- |
| *Acinetobacter baumannii* | Y | (14-16) |
| *Burkholderia spp* | Y | (16) |
| *Burkholderia cepacia* | Y | (16) |
| *Citrobacter fruendii* | Y | (14, 15) |
| *Citrobacter koseri* | Y | (14, 15) |
| *Klebiella aerogenes* | Y | (1, 14-16) |
| *Enterobacter cloacae* | Y | (1, 14-16) |
| *Enterococcus faecium* | Only if predominant | Microbiological archives |
| *Enterococcus faecalis* | Only if predominant | Microbiological archives |
| *Escherichia coli* | Y | (1, 14-16) |
| *Fusobacterium necrophorum* | Microbiological archives | (16) |
| *Fusobacterium nucleatum* | Microbiological archives | (16) |
| *Haemophilus influenzae* | Y | (1, 14, 16) |
| *Hafnia alvei* | Y | Microbiological archives |
| *Klebsiella oxytoca* | Y | (1, 14-16) |
| *Klebsiella pneumoniae* | Y | (1, 14-16) |
| *Klebsiella variicola* | Y | (14-16) |
| *Legionella pneumophila* | Y | (14-16) |
| *Moraxella catarrhalis* | Y | (1, 14-16) |
| *Morganella morganii* | Y | (14-16) |
| *Proteus mirabilis* | Y | (1, 14-16) |
| *Pseudomonas aeruginosa* | Y | (1, 14-16) |
| *Serratia marcescens* | Y | (1, 14-16) |
| *Stenotrophomonas maltophilia* | Y | (1, 2, 16) |
| *Streptococcus agalactiae* | Y | (14-16) |
| *Streptococcus pneumoniae* |  | (2, 14-16) |
| *Streptococcus pyogenes* | Y | (1, 14-16) |
| *Streptococcus viridans group (S. anginosus, S. intermedius, S. constellatus)* | Y | Microbiological archives & (16) |
| *Staphylococcus aureus* | Y | (1, 14-16) |
| *Tropheryma whipplei* | Only if predominant | Microbiological archives |
| *Mycobacterium tuberculosis* | Y | Microbiological archives |
| Mycobacterium non-tuberculosis | Y | Microbiological archives |
| *Corynebacterium striatum* | Only if predominant | Microbiological archives |
| **Fungi** |  |  |
| *A. fumigatus* | Y | (16) |
| *A.niger* | Y | Microbiological archives |
| *A.flavus* | Y | Microbiological archives |
| *A. versicolor* | Y | Microbiological archives |
| *A. nidulans* | Y | Microbiological archives |
| *A. glaucus* | Y | Microbiological archives |
| *A. terreus* | Y | Microbiological archives |
| *A. clavatus* | Y | Microbiological archives |

*All organisms listed were also identified in the archives of reportable organisms by microbiological culture in the last 5 years in the clinical laboratory.

### Table S11. Analytical limit of detection of the RMg workflow determined using a culture-negative BAL sample

| **Sample** | **Replicate** | **Pathogen** | **Approx. number of pathogen cells (CFU)** | **Total reads (2hrs)** | **Human reads (2hrs)** | **Microbial reads** | **Classified reads* mapped to the organism** |
| --- | --- | --- | --- | --- | --- | --- | --- |
| SA 10^4^ | 1 | ***S. aureus*** | 10,000 | 28997 | 53 | 18171 | 1469 (8%) |
| SA 10^4^ | 2 |  | 10,000 | 5946 | 78 | 3986 | 772 (19.3%) |
| SA 10^4^ | 3 |  | 10,000 | 39907 | 138 | 24381 | 3150 (13%) |
| SA 10^3^ | 1 |  | 1,000 | 31442 | 201 | 19125 | 211 (1%) |
| SA 10^3^ | 2 |  | 1,000 | 22459 | 65 | 13394 | 156 (1%) |
| SA 10^3^ | 3 |  | 1,000 | 35387 | 157 | 20797 | 61 (0.3%) |
| KP 10^4^ | 1 | ***K. pneumoniae*** | 10,000 | 32092 | 17113 | 9713 | 1412 (15%) |
| KP 10^4^ | 2 |  | 10,000 | 55750 | 120 | 36067 | 3411 (10%) |
| KP 10^4^ | 3 |  | 10,000 | 49353 | 74 | 26585 | 1187 (5%) |
| KP 10^3^ | 1 |  | 1,000 | 33494 | 594 | 22096 | 410 (1.86%) |
| KP 10^3^ | 2 |  | 1,000 | 59246 | 629 | 37031 | 765 (2%) |
| KP 10^3^ | 3 |  | 1,000 | 43532 | 156 | 27095 | 460 (2%) |
| CA 10^3^ | 1 | ***C. albicans*** | 1,000 | 15623 | 547 | 8579 | 46 (0.5%) |
| CA 10^3^ | 2 |  | 1,000 | 968 | 144 | 414 | 10 (2%) |
| CA 10^3^ | 3 |  | 1,000 | 15975 | 1595 | 7916 | 118 (2%) |
| CA 10^2^ | 1 |  | 100 | 2808 | 1760 | 788 | 3 (0.4%) |
| CA 10^2^ | 2 |  | 100 | 75719 | 38280 | 11372 | 0 |
| CA 10^2^ | 3 |  | 100 | 1120 | 952 | 103 | 2 (2%) |

*The number of reads detected for all samples was above the pre-defined thresholds after 2hrs of sequencing .

### References

1.	Charalampous T, Alcolea-Medina A, Snell LB, Williams TGS, Batra R, Alder C, et al. Evaluating the potential for respiratory metagenomics to improve treatment of secondary infection and detection of nosocomial transmission on expanded COVID-19 intensive care units. Genome Medicine. 2021;13(1):182.

2.	Charalampous T, Kay GL, Richardson H, Aydin A, Baldan R, Jeanes C, et al. Nanopore metagenomics enables rapid clinical diagnosis of bacterial lower respiratory infection. Nature biotechnology. 2019;37(7):783-92.

3.	Baldan R, Cliff PR, Burns S, Medina A, Smith GC, Batra R, et al. Development and evaluation of a nanopore 16S rRNA gene sequencing service for same day targeted treatment of bacterial respiratory infection in the intensive care unit. Journal of Infection. 2021;83(2):167-74.

4.	Services M. UK Standards for Microbiology Investigations. Evaluations, validations and verifications of diagnostic tests.

5.	Unit MS. UK Standards for Microbiology Investigations. Investigation of bronchoalveolar lavage, sputum and associated specimens.

6.	Justin Joseph O'grady GLK, Themoula CHARALAMPOUS, Alp AYDIN, Riccardo SCOTTI, inventorMethod for digesting nucleic acid in a sample. United Kingdom patent WO2021105659A1. 2109 03/06/2021.

7.	Kim D, Song L, Breitwieser FP, Salzberg SL. Centrifuge: rapid and sensitive classification of metagenomic sequences. Genome research. 2016;26(12):1721-9.

8.	Li H. Minimap2: pairwise alignment for nucleotide sequences. Bioinformatics. 2018;34(18):3094-100.

9.	Sichtig H, Minogue T, Yan Y, Stefan C, Hall A, Tallon L, et al. FDA-ARGOS is a database with public quality-controlled reference genomes for diagnostic use and regulatory science. Nature Communications. 2019;10(1):3313.

10.	O'Leary NA, Wright MW, Brister JR, Ciufo S, Haddad D, McVeigh R, et al. Reference sequence (RefSeq) database at NCBI: current status, taxonomic expansion, and functional annotation. Nucleic acids research. 2016;44(D1):D733-45.

11.	Seeman T. Abricate. Github. Available from: <https://github.com/tseemann/abricate>

12.	Andrew J Page TLV, Justin O'Grady. Scagaire. GitHub. 2019.

13.	Alcock BP, Raphenya AR, Lau TTY, Tsang KK, Bouchard M, Edalatmand A, et al. CARD 2020: antibiotic resistome surveillance with the comprehensive antibiotic resistance database. Nucleic acids research. 2020;48(D1):D517-d25.

14.	Gadsby NJ, McHugh MP, Forbes C, MacKenzie L, Hamilton SKD, Griffith DM, et al. Comparison of Unyvero P55 Pneumonia Cartridge, in-house PCR and culture for the identification of respiratory pathogens and antibiotic resistance in bronchoalveolar lavage fluids in the critical care setting. European journal of clinical microbiology & infectious diseases : official publication of the European Society of Clinical Microbiology. 2019;38(6):1171-8.

15.	Enne VI, Aydin A, Baldan R, Owen DR, Richardson H, Ricciardi F, et al. Multicentre evaluation of two multiplex PCR platforms for the rapid microbiological investigation of nosocomial pneumonia in UK ICUs: the INHALE WP1 study. Thorax. 2022;77(12):1220.

16.	Langelier C, Kalantar KL, Moazed F, Wilson MR, Crawford ED, Deiss T, et al. Integrating host response and unbiased microbe detection for lower respiratory tract infection diagnosis in critically ill adults. Proceedings of the National Academy of Sciences of the United States of America. 2018;115(52):E12353-e62.

17.	Pendleton KM, Huffnagle GB, Dickson RP. The significance of Candida in the human respiratory tract: our evolving understanding. Pathogens and disease. 2017;75(3).
