## Supplementary Figure S1 for "Routine respiratory metagenomics service for intensive care unit patients"

TP - True Positive
TN - True Negative
FN - False Negative
FP - False Positive
GPB - Gram Positive Bacteria
GNB - Gram Negative Bacteria
URTF - Upper respiratory tract flora

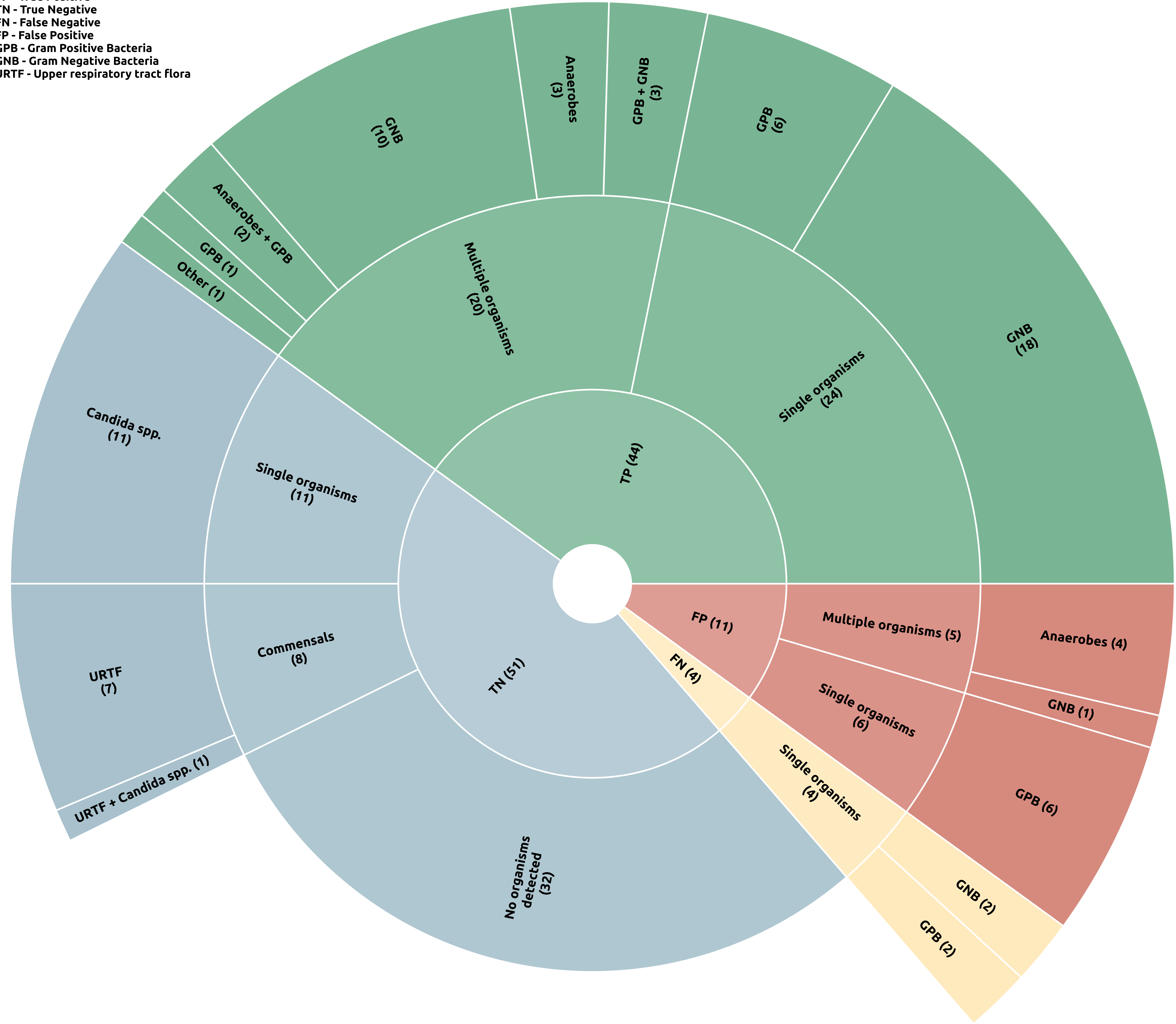
